# The “Great Lockdown”: Inactive Workers and Mortality by Covid-19

**DOI:** 10.1101/2020.09.17.20190595

**Authors:** Nicola Borri, Francesco Drago, Chiara Santantonio, Francesco Sobbrio

**Author notes:** We are also grateful to Alberto Bisin, Daniela Iorio and Simona Grassi for useful comments. We also thank Ruben Durante and Elliot Gaston Motte for their help in gathering the EnelX mobility data and Facebook which provided data on human mobility through its “Data for Good” program.

## Abstract

In response to the Covid-19 outbreak the Italian Government imposed an economic lockdown on March 22, 2020 and ordered the closing of all non-essential economic activities. This paper estimates the causal effects of this measure on mortality by Covid-19 and on mobility patterns. The identification of the causal effects exploits the variation in the active population across municipalities induced by the economic lockdown. The difference-in-differences empirical design compares outcomes in municipalities above and below the median variation in the share of active population before and after the lockdown within a province, also controlling for municipality-specific dynamics, daily-shocks at the provincial level and municipal unobserved characteristics. Our results show that the intensity of the economic lockdown is associated with a statistically significant reduction in mortality by Covid-19 and, in particular, for age groups between 40-64 and older (with larger and more significant effects for individuals above 50). Back of the envelope calculations indicate that 4,793 deaths were avoided, in the 26 days between April 5 to April 30, in the 3,518 municipalities which experienced a more intense lockdown. Several robustness checks corroborate our empirical findings.

## 1 Introduction

The immediate effects of the Covid-19 pandemic in Italy have been dramatic. First, and foremost, between February 2020 and April 2021, according to official reports, almost 120,000 people have died and around 3.6 million were infected. Second, the lockdown measures, which froze large parts of the economy, determined a large drop in economic activity with severe consequences for workers, firms, and public finances.

This paper uses a policy induced variation in active population across Italian municipalities after the economic lockdown in the spring 2020 to evaluate its effectiveness in controlling the pandemic at the beginning of the epidemic. Specifically, with our analysis, we try to estimate the causal effect of the intensity of the lockdown measures in reducing deaths by Covid-19. The main finding is that the intensity of the lockdown was significantly related to a reduction in mortality by Covid-19, and to a reduction in people’s mobility. Because the empirical strategy quantifies the effect of a reduction in the active population on deaths by Covid-19, our results are useful to discipline the parameters in epidemiology models of the diffusion, and containment, of a pandemic.

The analysis of the effects of the lockdown is important for at least two reasons. First, to understand the overall cost-effectiveness of the lockdown measures at the peak of the first wave of the pandemic (Chilton *et al*., 2020). Second, to guide and inform policy makers in the design of the social distancing measures in the event of a new wave of the pandemic. In fact, because “non-pharmaceutical interventions” (NPIs) differ from each other in terms of their economic (Bartik *et al*., 2020) and psychological (Brooks *et al*., 2020) costs, it is crucial “to identify the interventions that most reduce transmission at the lowest economic and psychological cost” (Haushofer and Metcalf, 2020).

In response to the Covid-19 outbreak, the Italian Government imposed a first lockdown on March 11, 2020 which closed many business activities open to the public like restaurants and gyms, and a second — economic — lockdown on March 22, which ordered the closing of all non-essential economic activities and prohibited any movement of people with few exceptions, like proven work or health related reasons.^1^ This paper focuses on the closing of non-essential economic activities of the second lockdown and, specifically, on the induced reduction in the share of active population across different municipalities – measured by the number of employed workers (15 years old and above) active in economic sectors not subject to the lockdown over the total population – to evaluate its impact on the spread of the Covid-19 pandemic and on mobility patterns.

The key outcome variable of our empirical analysis is the municipal daily-mortality by Covid-19, measured as the difference between the daily number of deaths in 2020 and the average number of deaths on the same day and in the same municipality between 2015 and 2019. In particular, we use official municipal-level data from the Italian National Statistical Institute (ISTAT) on the daily deaths from 2015 until April 2020. Following the literature and public health authorities, we consider excess deaths as a proxy of mortality by Covid-19 (and, implicitly, contagion) to overcome, at least partially, issues related to differences in the classifications of deaths due to Covid-19, testing, and hospital capacity (see, among others, Galeotti and Surico 2020; National Center for Health Statistics 2020; Woolf *et al*. 2020).^2^ Indeed, as pointed out by Woolf *et al*. (2020): “The number of publicly reported deaths from coronavirus disease 2019 (COVID-19) may underestimate the pandemic’s death toll. Such estimates rely on provisional data that are often incomplete and may omit undocumented deaths from COVID-19.” (Woolf *et al*. 2020, page 510).

Our empirical analysis exploits the geographical heterogeneity in the reduction of active population across Italian municipalities induced by the design of the economic lockdown. This heterogeneity derives from the combination of the share of active population in a municipality and the share of those working in a sector closed by the lockdown. Importantly, unlike the first lockdown, relevant heterogeneities in the share of active population emerged as a consequence of the economic lockdown even when comparing municipalities in the same province. This heterogeneity is also rather granular as our sample covers 7,089 municipalities with a mean (median) population of 7,729 (2,443) residents and a mean (median) area of 36 (21) squared kilometers, each belonging to one of the 110 Italian provinces. As such, the Italian economic lockdown provides a design to elicit the causal effect of the containment policy (Goodman-Bacon and Marcus, 2020). The identification of the causal effect uses a difference-in-differences design comparing excess deaths and mobility outcomes in municipalities above and below the median variation in the share of active population before and after the lockdown within a province, also controlling for municipality-specific dynamics, daily-shocks at the provincial level and municipal unobserved characteristics.

Our results show that the intensity of the economic lockdown is associated with a statistically significant reduction in excess deaths for the entire population and, in particular, for age groups between 40-64 and older (with larger and more significant effects for individuals above 50), consistent with the evidence that Covid-19 is particularly risky for older people (Dowd *et al*., 2020). We also document that the effects are similar and significant also when looking at males and females separately. A back of the envelope calculation indicates that, overall, in the 26 days between April 5 and April 30, in the 3,518 municipalities with a more intense lockdown, 4,793 deaths were avoided or 1.36 lives per municipality. In these municipalities the share of active population dropped on average by 42.5 percentage points compared to 17 percentage points in municipalities with a less severe lockdown. In terms of elasticity, and assuming linearity, our calculations suggest that a 1 percentage point reduction in the share of active population caused a 1.32 percentage points reduction in mortality by Covid-19 (as measured by excess deaths). We then turn to the analysis of the evolution of mobility around the first and second lockdowns in the municipalities above and below the provincial median drop in the share of active population. Although we find that the two groups are different in the pre-lockdown period, we also find that the group experiencing a stronger reduction of active population is also characterized by a stronger reduction in mobility in the second (economic) lockdown.

We ran a battery of checks to evaluate the robustness of our results. First, we show that our results are robust to an alternative source for the share of active population. Second, we show that our results are robust to using weighs for the share of active population by indices for physical proximity and propensity to work-from-home for each macro-sector, which allows us to take into account the possibility of some categories converting to working remotely and for differences in the availability of home working across sectors. Next, we consider a placebo exercise in which the treatment period is restricted to the two weeks after the beginning of the second lockdown. In this case, we find a smaller and not significant coefficient associated with the effect of the lockdown, which is consistent with the fact that our results are not driven by pre-trends. We also show that our results are robust to the use of alternative definitions of intensity of the lockdown. Importantly, we obtain similar results with a linear model (i.e., assessing the impact of a linear drop in the share of active population on excess deaths). In addition, we show that our results also hold when using different percentiles (rather than the median) as cutoffs for identifying municipalities experiencing a more intense lockdown. Finally, we restrict the sample to municipalities below several thresholds of population size, or according to narrow margins of variation in the reduction of active population, and we compare again municipalities with more and less intense lockdowns. Also in these cases we find a significant effect.

This paper contributes to the large and growing economic literature spurred by the Covid-19 pandemic. One strand of the literature studies theoretical models that extend the classic SIR framework of Kermack and McKendrick (1927) to account for province-level spatial networks (Gatto *et al*., 2020); for the interaction between economic decisions and epidemics (Eichenbaum *et al*., 2020); for the optimal lockdown policy accounting for the trade-off between economic cost and fatalities (Alvarez *et al*., forthcoming); and for multiple regions (countries, states and cities) to infer unobservables, like the number of recovered (Fernández-Villaverde and Jones, 2020). With respect to this strand of the literature, the results of our paper are useful to discipline the parameters of the theoretical models and to simulate the effects of alternative lockdown measures (see for example Favero *et al*. (2020)). To design effective policies to reduce the spread of the epidemics, scholars should also incorporate economic agents’s behavioral responses in epidemiological models (Bisin and Moro, 2020; Briscese *et al*., 2020; Jamison *et al*., 2020; Sheridan *et al*., 2020). Our paper, using detailed data on mobility, contributes to estimate such behavioral responses. More broadly, our paper is related to the literature assessing the possible socio-economic drivers of the Covid-19 pandemic. In terms of scope and methodology, our paper is closer to the literature that empirically studies the effects and effectiveness of lockdown and social distancing measures, mostly using natural experiments created by heterogeneity in policies. Adda (2016), using a sample that spans twenty five years before the Covid-19 pandemic, finds that the effectiveness of social distancing and lockdown measures depends on the capacity to reduce people mobility. Lyu and Wehby (2020b) find an increase in rates of Covid-19 cases in border counties in Iowa compared with border counties in Illinois after a stay-at-home order was implemented in Illinois but not in Iowa; Fang *et al*. (2020) and VoPham *et al*. (2020) find significant reductions in the diffusion of the pandemic, respectively in China and in the US, associated with a stronger drop in mobility and increase in social distancing. The paper closest to ours is Glaeser *et al*. (2020) which estimates the effect of mobility reduction on Covid-19 contagion at the zip-code level in five large US cities, instrumenting mobility with the share of active workers. We complement their findings by providing evidence from Italy, one of the first country hit by the pandemic. Importantly, with respect to the existing literature, our paper looks at excess deaths which are not influenced by (possibly endogenous) testing policies, classifications of deaths due to Covid-19 and hospital capacity. Last but not least, our analysis employs rather granular panel data by exploiting municipality-day variations in the share of active population as a result of the heterogeneous impact of lockdown policies in Italy. By looking at within province-day variation in excess deaths, while also controlling for municipal fixed effects and lagged municipal excess-deaths, our study compares treated and control municipalities that are plausibly homogenous from an ex-ante perspective both in terms of contagion dynamic and health care system.^3^

## 2 Data

The data used in this paper come from three sources. First, we construct a measure of daily “excess deaths” using official data from ISTAT, at the municipal level, for the period January 1 to April 30 of each year starting in 2015 and ending in 2020. Excess deaths are computed as the difference in the daily number of deaths at the municipal-day level in 2020 with respect to the municipal daily average in the years 2015 to 2019. As discussed in the introduction, we consider excess deaths as a proxy of mortality by Covid-19 (and implicitly of contagion) to overcome potential issues related to the endogeneity of testing policies, hospital capacity and death classification at the local level (Galeotti and Surico 2020; National Center for Health Statistics 2020; Woolf *et al*. 2020).^4^

Second, we exploit the geographical heterogeneity in the share of active population due to the selective and progressive restriction of sectors subject to the first lockdown (March 11, 2020) and to the second (economic) lockdown (March 22, 2020). In particular, as explained in details in the Online Appendix A, business activities open to the public were closed first, and later all non-essential economic activities. We combine data on the number of workers at the NACE 3-digit sector level with the list of sectors excluded by the lockdown to retrieve the time-varying fractions of inactive (and active) population at the municipal-daily level. Specifically, we compute the share of active population during the first lockdown as the number of active workers (15 years old and above) on March 11 (i.e., following the first lockdown restrictions) over the total population of the municipality. Analogously, we compute the share of active population during the second lockdown as the number of active workers on March 22 (i.e., following the second lockdown restrictions) over the total population of the municipality.^5^ Third, we construct two mobility indicators, also at the municipal-daily level, using data from City Analytics by EnelX and HERE Technologies (EnelX henceforth), which are based on anonymous aggregate data from connected vehicles, navigation maps and systems, such as geolocation data from mobile apps. For each municipality, the first indicator measures the distance in kilometers covered by individuals, while the second indicator measures their movements. Both measures are expressed as ratios with respect to the municipal population. Online Appendix C provides a validation of these two mobility measures by comparing them with an alternative indicator built using data provided by Facebook for a subset of municipalities.

## 3 Methods

Let *ED*_*ijt*_ denote the excess deaths of municipality *i* in province *j* at date (day) *t*. Exploiting the panel structure of the data and the heterogeneity in the severity of the second lockdown across Italian municipalities, the aim is to estimate the causal effect of the intensity of the lockdown on *ED*.

The first challenge is the measurement of the intensity of the lockdown that we denote with *L*. We can measure *L* with the reduction in mobility within each municipality or with any measure that captures the reduction of economic and social interactions. Such a measure, however, is endogenous to *ED* since a given reduction in economic and social activity might be the result of a behavioral response to ED. Furthermore, the reduction in mobility is potentially correlated with factors that have an impact on *ED*. For example, citizens might reduce economic and social activity more in municipalities in which the quality of hospital care is lower and in turn where *ED* is higher. To circumvent these problems we consider the share of active municipal population before and after the second lockdown, described in section 2, which is predetermined to Covid-19. For each province *j* we calculate the median reduction in the share of active municipal population from the first (March 11) to the second lockdown (*t*_0_=March 22). We assign *L*_*ijt*_=1 if *t ≥ t*_0_+14 and the municipality *i* is above the median reduction in the share of active population in its province *j*, and *L*_*ijt*_ = 0 in all other cases.^6^ The hypothesis, that we are able to test, is that the above-the-median municipalities experienced a more severe lockdown in terms of reduction in mobility. Following the medical literature (Lauer *et al*., 2020; McAloon *et al*., 2020; Wilson *et al*., 2020; Sun *et al*., 2020; Istituto Superiore di Sanit`a, 2020), to capture the consequences of the second lockdown on our *ED* measure, we impose that the potential effects on deaths start with a two-week gap. Indeed, as pointed out by Lauer *et al*. (2020) and McAloon *et al*. (2020), the median incubation period for Covid-19 (i.e., from the initial contagion to the onset of symptoms) is around 5 days. Moreover, according to the Italian Istituto Superiore di Sanit`a (2020) (the leading technical-scientific body of the Italian National Health Service) the median time between the onset of symptoms and death, for Covid-19 patients in Italy, was 12 days. This implies that half of the deaths observed over our sample period are likely to occur within 17 days from the initial contagion. Therefore, by imposing a 14-days gap in the start of our treatment period following the economic lockdown, we are adopting a rather conservative approach likely leading to underestimate the effect of the economic lockdown. Furthermore, in the empirical analysis we show that — indeed — the economic lockdown had no statistically significant effect on excess deaths reduction when looking at only the first two weeks after March 22.

In Figure 1 we plot the evolution in the 5-day moving average of *ED* for the two groups of municipalities. The Figure shows that—despite the closing of schools and the other containment measures of the first lockdown—both groups of municipalities experienced a rise in *ED* at the beginning of March which lasted until the end of the month. Importantly, above-the-median municipalities experienced a sharper *increase* in excess deaths in the first lockdown period (March 11-March 22) while they experienced a sharper decrease in the second lockdown period (March 22-April 30) compared with below-the-median ones. In line with a mechanism linking active population and mortality by Covid-19, this is suggestive evidence that municipalities characterized by a higher share of active population paid a higher death toll before the second (economic) lockdown, while they experienced a more marked reduction of mortality by Covid-19 once the lockdown was in place. Although part of the reduction in excess death after March 22 was plausibly a consequence of the first lockdown, it is not obvious why the latter should have had a differential effect on municipalities above/below the median drop in the share of active population. In fact, the intensity of the first lockdown, which imposed the closure of schools and some business activities open to the public, was substantially similar across municipalities. Indeed, the median drop in the share of active population due to the second lockdown (0.237) was more than 6.5 times larger than the median drop due to the first lockdown (0.036). Similarly, the induced cross-sectional variation in the share of active population due to the first lockdown was rather limited both in absolute and relative terms compared with the second lockdown (the standard deviation of the drop in the share of active population across municipalities was 0.048 in the first lockdown, and 0.239 in the second lockdown). It is, thus, less plausible that the differential reduction in excess deaths after April 5 (i.e., the start of our treatment period) depends on the effect of the first lockdown, which occurred almost a month before and affected in a very similar way all municipalities. Table 1 reports summary statistics for the average *ED* reported in Figure 1 and the share of active population for the two groups of municipalities before and after the second lockdown. It is important to remark that by considering municipalities above vs. below the provincial median drop in the share of active, we can more transparently compare pre-trends in municipalities differentially affected by the lockdown.^7^

**Table 1:** Summary Statistics

|  | Below-the-Median<br>Municipalities | Above-the-Median<br>Municipalities |
| --- | --- | --- |
| **Control Period**<br>(March 11 - April 4) |  |  |
| Total excess deaths | 7062 | 21358 |
| Daily excess deaths (ED) | 0.0791<br>(0.521) | 0.243<br>(1.545) |
| Share of active population* | 0.366<br>(0.221) | 0.831<br>(0.514) |
| **Treatment Period**<br>(April 5 - April 30) |  |  |
| Total excess deaths | 2790 | 9484 |
| Daily excess deaths (ED) | 0.0301<br>(0.409) | 0.104<br>(1.044) |
| Share of active population | 0.196<br>(0.156) | 0.406<br>(0.287) |
| Municipalities | 3,571 | 3,518 |

Notes. The Table reports the total deaths in excess in the period (rounded to the closest integer), the mean daily excess deaths and the mean share of active population. The standard deviations for the daily excess deaths and share of active population are reported in parentheses. \*The share of active population for the control period (i.e., March 11-April 4) is reported as the one occurring in the first lockdown period (March 11 - March 22).

**Figure 1:**
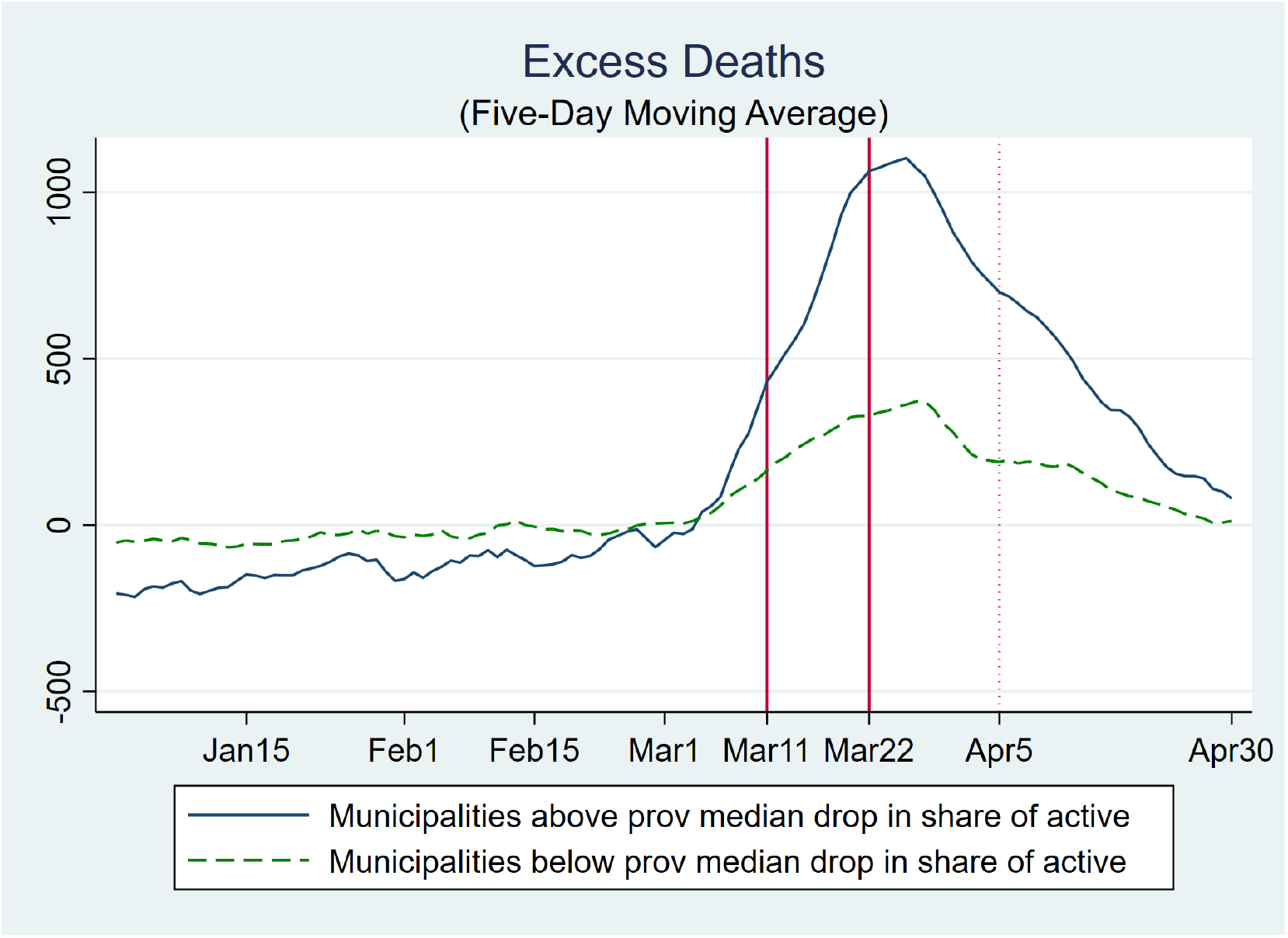
Excess Deaths (2020 vs 2015-2019) Notes. The figure illustrates the evolution in the 5-day moving average of excess deaths in the municipalities above (below) the median with respect to the drop in the share of active population in their province between the first and second lockdowns (respectively, blue line and green-dashed line). The two vertical lines indicate the dates of the first (March 11) and second (March 22, economic) lockdown. The dotted vertical line indicates the end of the two-week gap after the economic lockdown (April 5).

This empirical setting is similar to a difference-in-differences design in which we compare municipalities above and below the median reduction of active population before and after April 5. That is, as pointed out before, our control period is the one corresponding to the first lockdown (i.e. starting in March 11), including its lagged effects up to two weeks after its ending (i.e. until April 5). In general, we can control for time-invariant municipality characteristics (municipality fixed effects) and for shocks that are province specific and time variant (province-by-date fixed effects). In our case, the parallel trend assumption (namely that in the absence of the second lockdown, within the same province, municipalities with *L*=0 and with *L*=1 would have had the same trend) is not supported *prima facie* graphically: while we cannot observe a scenario without the second lockdown, the fact that between March 11 and April 5 differences in *ED* for the two groups of municipalities are not constant over time does not lend credibility to the parallel trend assumption. However, including in our specification a set of lags in *ED* may control for the diverging dynamics of excess deaths. In this case, the identifying assumption is that municipalities above and below the median with same level of past *ED* are not on a different trend in *ED*, once we control for municipality and province-by-date fixed effects. The model that we estimate is the following:

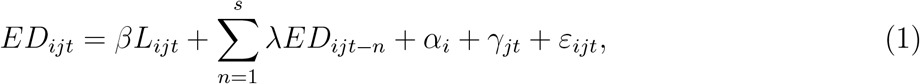

where in the main specification we fix *s*=7, the coefficient of interest is *β* (that we expect to be negative), and *α*_*i*_ and *γ*_*jt*_ are municipality and province-by-date fixed effects. Since we compare a period in which the first lockdown was implemented with a period associated with a more severe lockdown, Model 1 is estimated using observations from March 11 to April 30. The model requires a sequential exogeneity assumption. In other words, contemporaneous shocks on *ED* are allowed to have a feedback effect on future realizations of *ED* or be correlated with future values of *L*, but we assume that they cannot be correlated to past *ED* and with *L*. Formally, the assumption is that 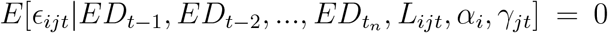 for all *t ≥ t*_*n*_. To fix ideas, this assumption requires that unobserved NPIs (e.g., further local mobility restrictions)— specific to a municipality and varying within province *j*—while having an effect on future and contemporaneous realizations of *ED*, are not correlated to past values of *ED* or with *L*. Since this is a strong assumption, we will also estimate Model 1 with fixed effects and with lagged dependent variables only. The estimation of Model 1 excluding the lagged values of *ED* or the set of fixed effects requires less restrictive assumptions and provides an upper and a lower bound of the causal effect of *L* on *ED*.

Indeed, by estimating Model 1 with fixed effects but without lags of *ED*, we obtain a coefficient (that we denote with 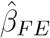) that is biased upward in absolute value: the model with fixed effects removes group specific characteristics and province-by-date shocks from 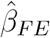, but ignores the fact that municipalities with a more severe lockdown experienced a sharper increase of *ED* before the lockdown. Hence, the diverging trends are incorporated in 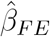 which is — in absolute value — higher than the “true” effect of *L* on *ED*. On the other hand, Model 1 with only lagged *ED* and a group indicator for the municipalities corrects for the diverging pre-trends but also absorbs declines in *ED* caused by the differential intensity of the lockdown, that otherwise would be incorporated in 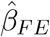. Hence, Model 1 with lagged *ED* provides an estimate of *L* on *ED* (that we denote with 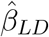) that is lower — in absolute value — than the “true” effect of *L* on *ED*. Because estimating Model 1 with only fixed effects or with only lagged *ED* gives us an upper and lower bound of the causal effect of *L* on *ED*, then a low difference between 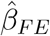 and 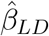 is consistent with the idea that there are no important violations of the identifying assumption leading to a large bias of 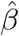 estimated in Model 1.^8^

## 4 Results

We focus on a set of municipalities (7,089) with complete deaths and active population data, accounting for 92.2% of the Italian population, in order to have a homogenous and balanced sample across all empirical specifications. Table 2 reports the results of estimating variations of Model 1 by different age groups and for the entire population. In the first column, in addition to our main variable (*L*), we include a group indicator for municipalities above and below the median and a dummy variable to control for the post-economic lockdown that is equal to one from April 5 onwards. In the second and third columns, we augment the latter specification with only lagged *ED* (reporting 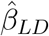) and with only fixed effects (reporting 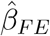), while the last column reports 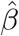 from Model 1. Table 2 illustrates two main patterns. First, the coefficient 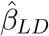 in Column 2 is lower in absolute value than the coefficient 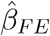 in Column 3, while (with only one exception) the coefficient in Column 4 lies in between the two coefficients. As discussed above, the coefficients reported in Column 2 and 3 provide an upper and lower bound of the true causal effect of the intensity of the lockdown proxied by *L*. Second, consistently with the fact that Covid-19 is particularly risky for older-age groups, we observe a statistically significant effect for the entire population and for age groups between 30-64 and older.^9^ Additional analysis by gender in Tables B.4 and B.5 (in Online Appendix B) also reveals that the lockdown seemed to have had similar effects on males and females across all age-groups with the exception of the 30-64 cohort showing more significant effects for males.

**Table 2:**
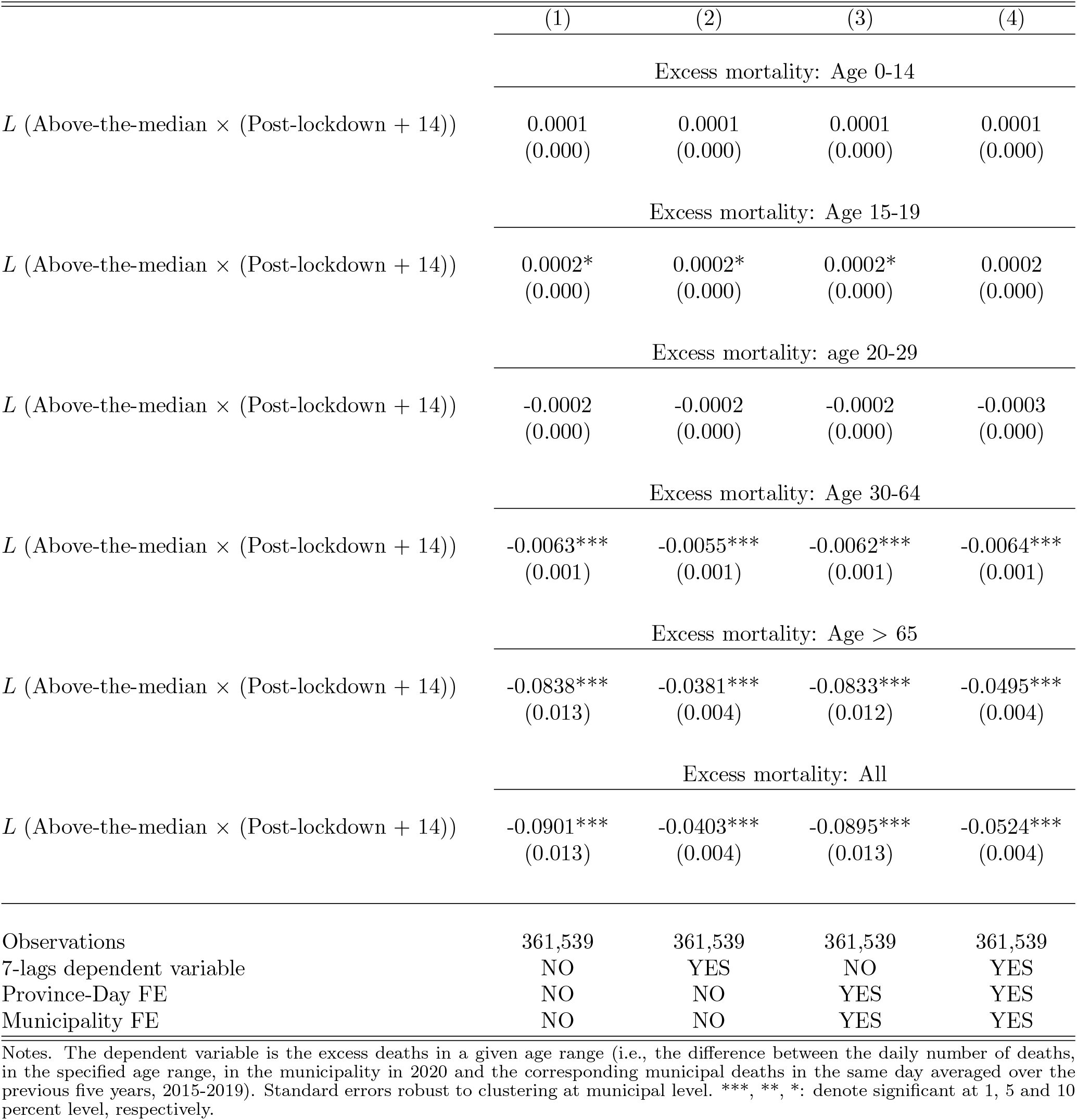
Economic Lockdown and Excess Deaths

Before discussing the magnitudes, robustness and limitations of the results in Table 2, we analyze how our measure of the intensity of the lockdown (*L*) is effectively correlated to our aggregate mobility measures. Figure 2 illustrates the evolution of aggregate mobility in municipalities above and below the median reduction in the share of active population by plotting the 5-day moving average of the daily kilometers per 1,000 residents.^10^ While the two groups are different in the pre-lockdown period (above-the-median municipalities on average have higher mobility), they both experience a sharp drop in mobility in the first-lockdown period. Some activities (e.g., schools and restaurants) closed in the first lockdown period and likely induced the observed reduction in mobility, but their overall impact in terms of contraction in active population—as already pointed out in Section 3—was rather limited and, importantly, rather homogeneous across municipalities compared with the second lockdown. Accordingly, as illustrated by Figure 2, in the second lockdown period, the “above-median” municipalities experienced a stronger reduction in mobility. That is, the economic lockdown caused an additional drop in mobility, likely driven by the number of workers employed in the sectors targeted by the lockdown.

**Figure 2:**
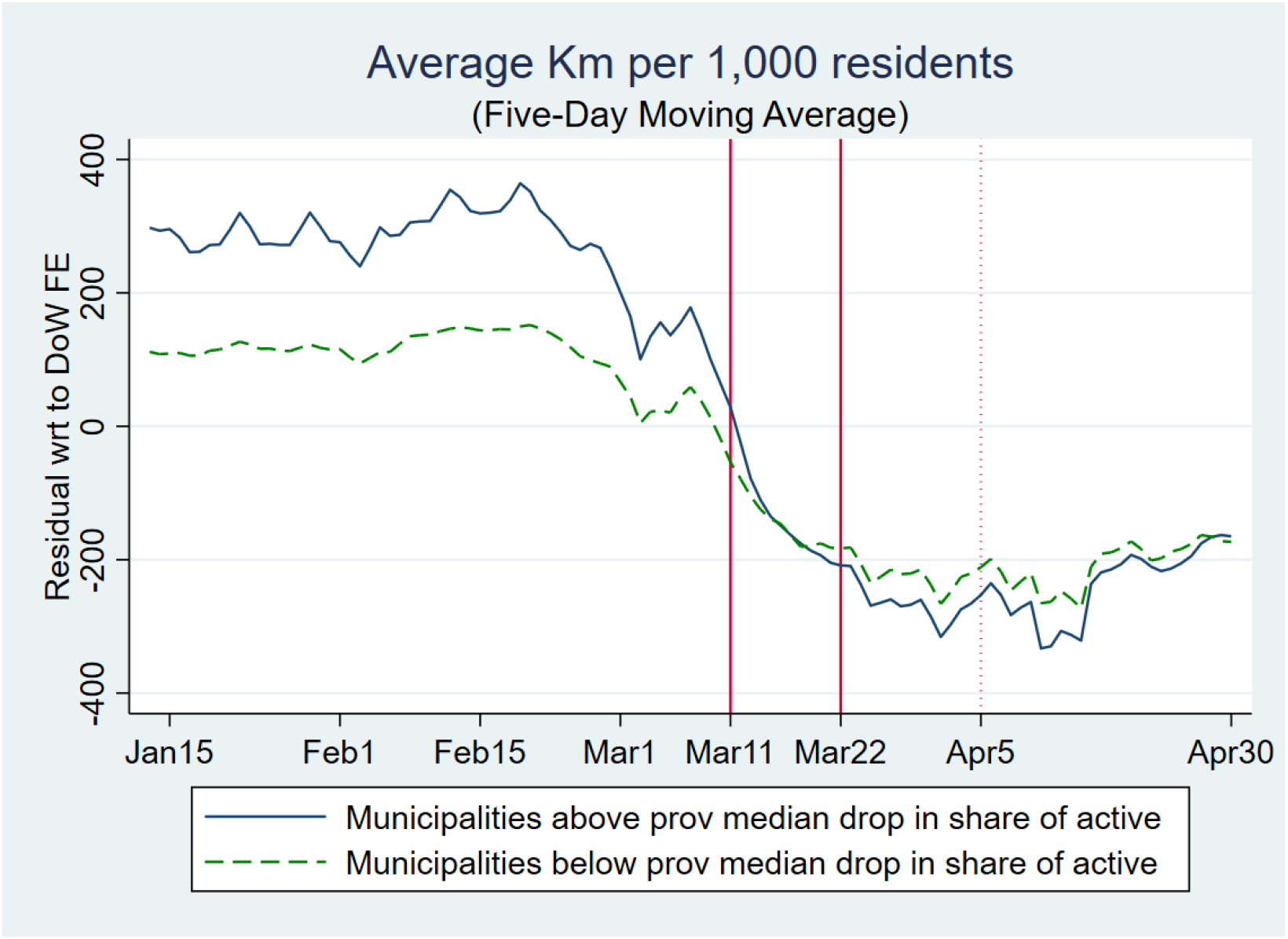
Kilometers per capita. Notes. The figure illustrates the evolution in the 5-day moving average in the kilometers per 1,000 residents (as residuals with respect to day-of-the-week fixed effects) in municipalities above (below) the median with respect to the drop in the share of active population the first and second lockdowns (respectively, blue line and green-dashed line). The two vertical lines indicate the dates of the first (March 11) and second (March 22, economic) lockdowns. The dotted vertical line indicates the end of two-week gap after the economic lockdown (April 5).

Table 3 presents the same evidence in a regression framework in which we regress our measure of mobility between March 11 and April 30 on our main indicator *L*_*ijt*_ and and a group indicator for municipalities above and below the median and a dummy variable to control for the post-economic lockdown (in Column 1) and a full set of municipality and province-by-date fixed effects (in Column 2). Unlike Model 1, we do not include lagged values of the dependent variable since we do not need to control for underlying contagion dynamics as for excess deaths because *L*_*ijt*_ is assumed to have an immediate effect on mobility. Hence, it is equal to 1 if *t ≥ t*_0_ (March 22) and the municipality *i* is above the median reduction in the share of active population in province *j*. Table 3 confirms the graphical intuition of Figure 2 and shows that it is robust to the inclusion of municipality and province-by-date fixed effects: after the second lockdown, municipalities with a larger contraction in the share of active population experienced a reduction in daily aggregate mobility of around 53 kilometers per 1,000 residents with respect to municipalities with a smaller contraction in the share of active population. Importantly, this reduction is more significant (both statistically and in terms of magnitude) in working days (Monday-Friday) relative to the weekends (Saturday-Sunday). Thus, our key indicator of reduction in economic activity is associated with a substantial reduction in aggregate mobility and particularly so in weekdays. Besides providing evidence on the effect of the economic lockdown on mobility, Table 3 also provides a validation of our lockdown measure (*L*) based on the difference between the share of active population between the second (economic) and first lockdowns. Online Appendix C shows that analogous results hold when considering an alternative measure of mobility based on the number of movements per 1,000 residents. Furthermore, we also show that the evidence presented in Figure 2 and Table 3 is consistent with the analysis using different mobility data from Facebook. It is important to remark that our mobility measures are aggregates at the municipality-daily level. As such, while they might provide useful insights on the impact of lockdown measures on aggregate human mobility at the municipal level, they are not able to capture possible underlying heterogeneous effects of social interactions among different population groups (e.g., school age vs. working age groups, see Section 5 for a discussion on this issue).

**Table 3:**
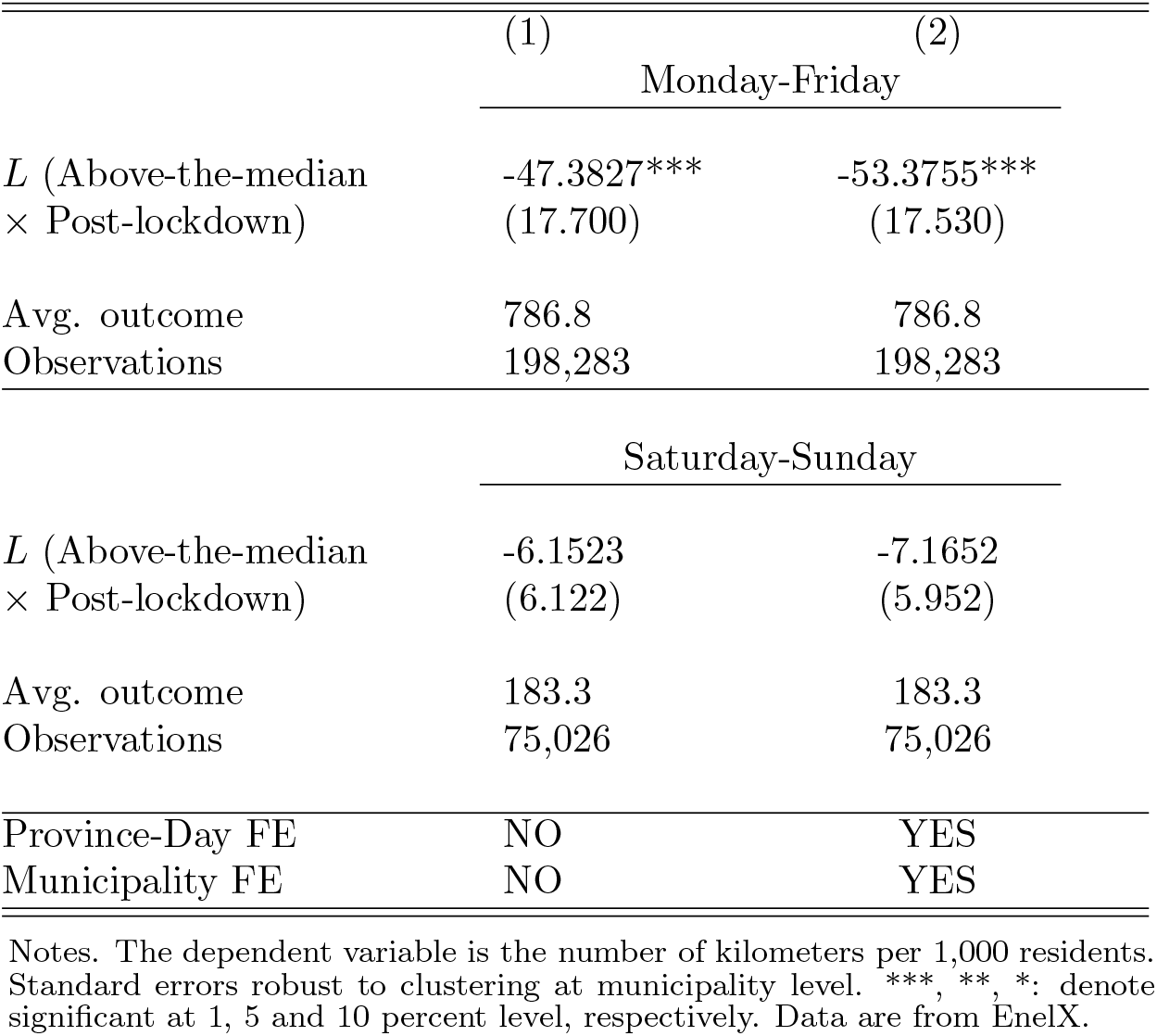
Economic Lockdown and Mobility

### 4.1 Magnitudes

Table 2 (Column 4) shows that being above the median reduction in the share of active population in its province after the lockdown reduces the difference in the excess deaths with respect to municipalities below the median by approximately 0.05. Looking at Table 1, this represents a 1/3 reduction of the difference in excess mortality between the two groups of municipalities in the pre-lockdown period (*≈* 0.05*/*(0.24 *−* 0.08)), i.e., before March 22 plus the two-week gap. For the municipalities that were hit severely by the second lockdown (the above-the-median), the total number of excess deaths in the April 5-30 period was 9,484. As back of the envelope calculation, consider a scenario in which the above-the-median municipalities had the same reduction in active population of the below-the-median municipalities (i.e., that above-the-median municipalities had the same lockdown intensity of below-the-median municipalities). In this scenario, the 3,518 above-the-median municipalities would have had approximately 4,793 additional excess deaths in the 26 days between April 5 and April 30 (i.e., 0.0524 *×* 26 *×* 3, 518 *≈* 4, 793). This represents, in our sample, an average of 1.36 lives saved per municipality.

To interpret these estimates note that above-the-median municipalities experienced an average reduction of the share of active population of 42.5 percentage points. By contrast below-the-median municipalities experienced an average reduction of the share of active population of 17 percentage points. Thus, increasing the intensity of the lockdown, measured by a reduction of the share of active population of 25.5 percentage points, reduced excess deaths by 33.6 percent (i.e. 9484*/*(9484 + 4793) *−* 1). Under the assumption that the effect is linear, the elasticity of mortality by Covid-19 (measured as excess deaths) with respect to a 1 percentage point reduction in the share of active population is 1.32. It should be clear that this elasticity applies to the Italian economic lockdown that was preceded by the first lockdown and the closure of schools, universities, and sport events (see Online Appendix A).

### 4.2 Robustness

Table 4 provides several robustness checks corroborating our results. The first column presents results by using an alternative source of data for the share of active population in the second (economic) lockdown. In particular, the provincial median (in the drop in the share of active population) is calculated by using data on the number of active workers in the second lockdown (calculated at the 5-digit NACE sectors) provided by ISTAT.^11^ In Columns 2 and 3, the shares of active population in the first and second lockdown are weighted by a physical proximity index and an inverse work-from-home (WFH) index of each NACE macro-sector (Barbieri *et al*., 2020). These specifications allow to take into account the characteristics of different economic sectors with respect to risk-exposure and in particular the possibility of some categories switching to remote working.^12^ Column 4 presents a robustness exercise where we drop the first two weeks of the control period (March 11-March 25) to account for possible lag effects of the first lockdown manifesting 14 days from its beginning (i.e., March 11). Finally, Column 5 presents a placebo exercise where the treatment period is restricted to the two weeks after the beginning of the economic lockdown, that is, comparing the first lockdown period with the 14 days following the beginning of the second lockdown period. We find that the coefficient is smaller than in our baseline specification and not statistically significant. As discussed above, absent pre-trends correlated with future realizations of *ED*, we should not expect an immediate significant effect of the economic lockdown on Covid-19 deaths. This placebo exercise seems to confirm that indeed this is the case.

**Table 4:**
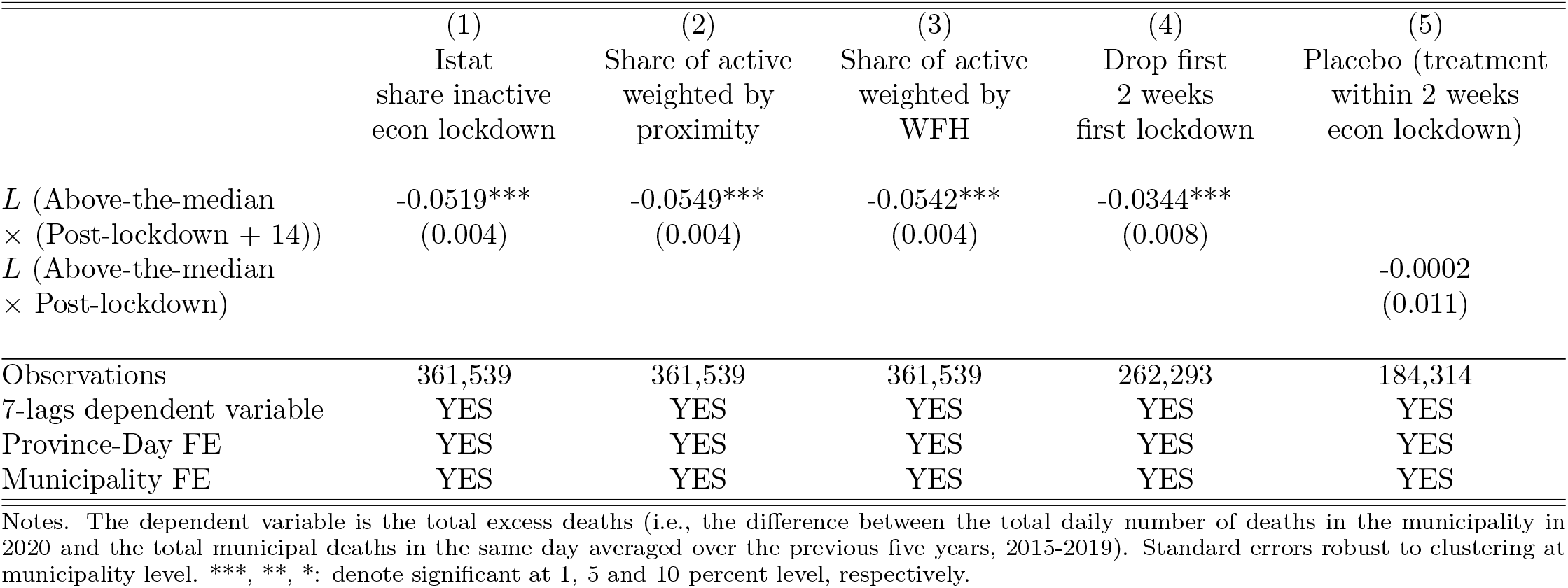
Economic Lockdown and Excess Deaths: Robustness

Online Appendix B provides further evidence that our results are robust with respect to different alternatives: *i*) categorize the municipalities according to the median variation of active workers in a province (rather than the median variation of active population) (Table B.1); *ii*) use of alternative thresholds in order to categorize municipalities as treated, i.e. to build our dummy *L* (Table B.2); *iii*) further breaking down the 30-64 age cohort (Table B.3) ; *iv*) looking at the male/female sub-groups (Tables B.4 and B.5) ; *v*) considering several additional specifications and extensions (Table B.6)

Finally, Table 5 presents results when considering the linear (time-varying) share of active population that is equal to the the share of active population of the first lockdown in the control period March 11-April 4, and to the share of active population of the second lockdown in the treatment period April 5-April 30. Furthermore, analogously to their counterparts in Table 4, Column 2 and 3 present results also for the share of active population weighted by a physical proximity index and an inverse work-from-home (WFH) index of each NACE macro-sector. Consistently with our baseline model, the linear specification shows that total excess death is positively associated with a higher share of active population. The coefficient for the specification where the share of active population is weighted by an inverse work-from-home (WFH) index is smaller than the unweighted one, which might suggest that some sectors probably switched to WFH prior to the legal lockdown.

**Table 5:** Economic Lockdown and Excess Deaths: Linear Model

|  | (1) | (2) | (3) |
| --- | --- | --- | --- |
| Share of active population | 0.0915***<br>(0.018) |  |  |
| Share of active population<br>weighted by proximity |  | 0.1112***<br>(0.022) |  |
| Share of active<br>weighted by WFH |  |  | 0.0598***<br>(0.012) |
| Observations | 361,539 | 361,539 | 361,539 |
| 7-lags dependent variable | YES | YES | YES |
| Province-Day FE | YES | YES | YES |
| Municipality FE | YES | YES | YES |

Notes. The dependent variable is the total excess deaths (i.e., the difference between the total daily number of deaths in each municipality in 2020 and the total municipal deaths in the same day averaged over the previous five years, 2015-2019). Standard errors robust to clustering at municipality level. \*\*\*, \*\*, \*: denote significant at 1, 5 and 10 percent level, respectively.

## 5 Discussion

There are two main alternative explanations for the baseline results presented in Tables 2, 4 and 5. The first is a reversion-to-the mean argument, the second is related to the effect of the first lockdown on mobility.

As illustrated by Figure 1, above-the-median municipalities had an increasing trend of excess deaths before April 5, compared to other municipalities. According to the reversion-to-the-mean explanation, these municipalities would have experienced a lower trend after April 5 even without a lockdown. We present three arguments not consistent with this explanation. First, the inclusion of the lags of the dependent variable controls for the dynamics of *ED*. The fact that we still find a substantial effect in Column 2 of Table 2, similar to that of Column 4, indicates that reversion to the mean cannot fully account for our results. Second, if the reversion-to-the-mean was a key driver, we should observe it just after the peak in excess deaths or around March 22. However, our placebo exercise (Column 5 in Table 4) tends to exclude this hypothesis. Third, when we narrow the sample around the provincial median drop in the share of active population – by analyzing subsamples of municipalities in 20-80, 30-70 and 40-60 percentiles in the reduction of the active population – we obtain groups of municipalities above and below the median reduction with much less diverging, or roughly parallel, trends before the lockdown (see Figure 3). In all these cases, the necessity to control for the lagged *EM* is less compelling. In fact, the fixed effects specification still identifies an effect of *L* on *ED* (see Table B.7 in Online Appendix B). The fact that we find an effect also in sub-samples where reversion-to-the-mean is not a concern (significant for the 20-80 and 30-70 percentiles) indicates that our main results are not due to differential pre-trends. Similarly, when looking at different sub-samples of municipalities below several population thresholds, we compare group of municipalities with very similar pre-trends in terms of excess-deaths, as illustrated by Figure 4, and still find an effect (see Table B.8 in Online Appendix B).

**Figure 3:**
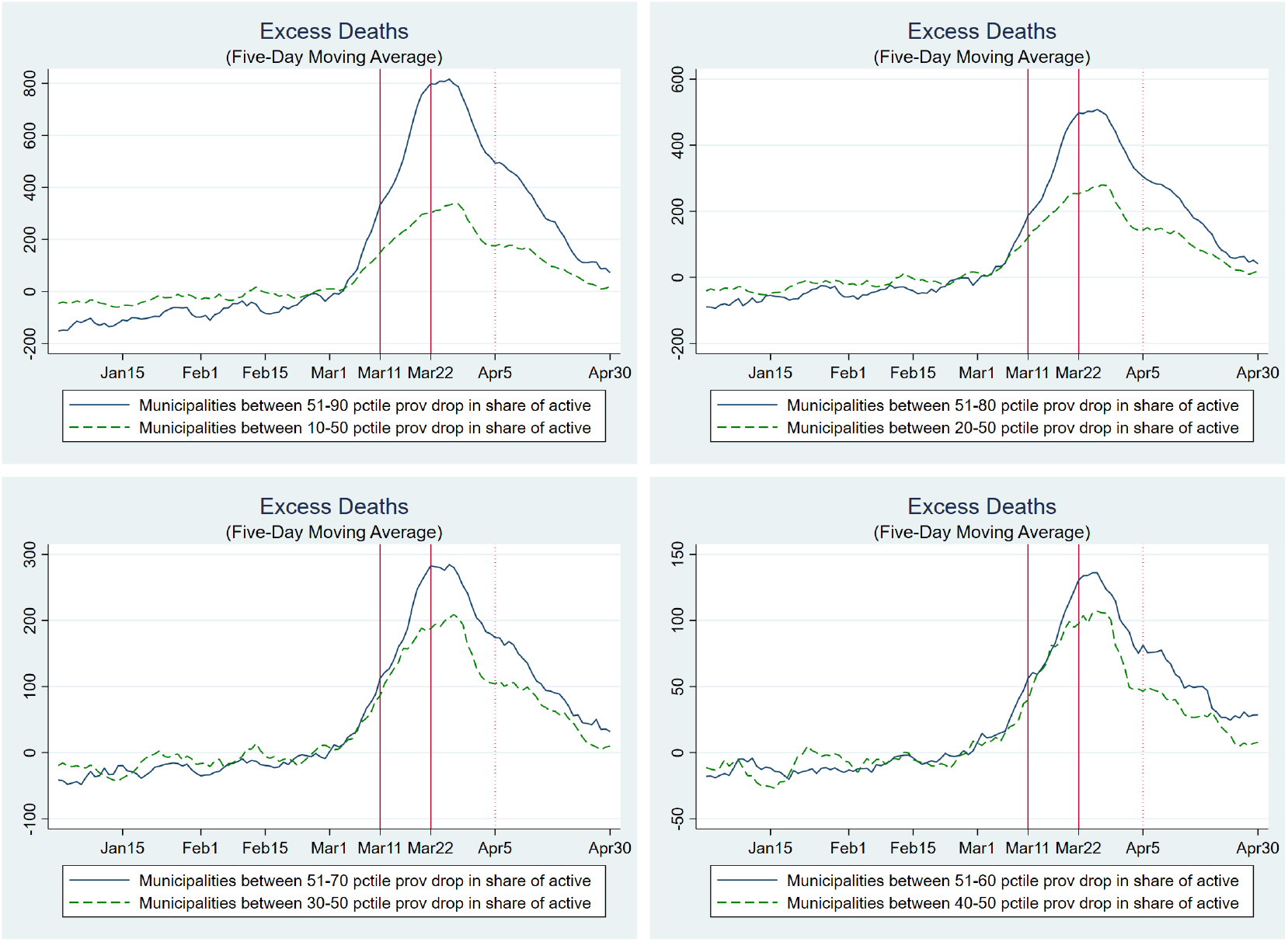
Excess Deaths: Subsamples by percentile drop share of active in the province. Notes. The figure illustrates the evolution in the 5-day moving average of excess deaths at the municipal level in the treated group (blue line) and control group (green-dashed line) in different sub-sample. In the top-left panel, the treated (control) group contains the municipalities above (below) the median and below (above) the 90th (10th) percentile with respect to the drop in the share of active population in their province between the first and second lockdowns. In the top-right panel, the treated (control) group contains the municipalities above (below) the median and below (above) the 80th (20th) percentile with respect to the drop in the share of active population in their province between the first and second lockdowns. In the bottom-left panel, the treated (control) group contains the municipalities above (below) the median and below (above) the 70th (30th) percentile with respect to the drop in the share of active population in their province between the first and second lockdowns. In the bottom-right panel, the treated (control) group contains the municipalities above (below) the median and below (above) the 60th (40th) percentile with respect to the drop in the share of active population in their province between the first and second lockdowns. The two vertical lines indicate the dates of the first (March 11) and second (March 22, economic) lockdown. The dotted vertical line indicates the end of the two-week gap after the economic lockdown (April 5).

**Figure 4:**
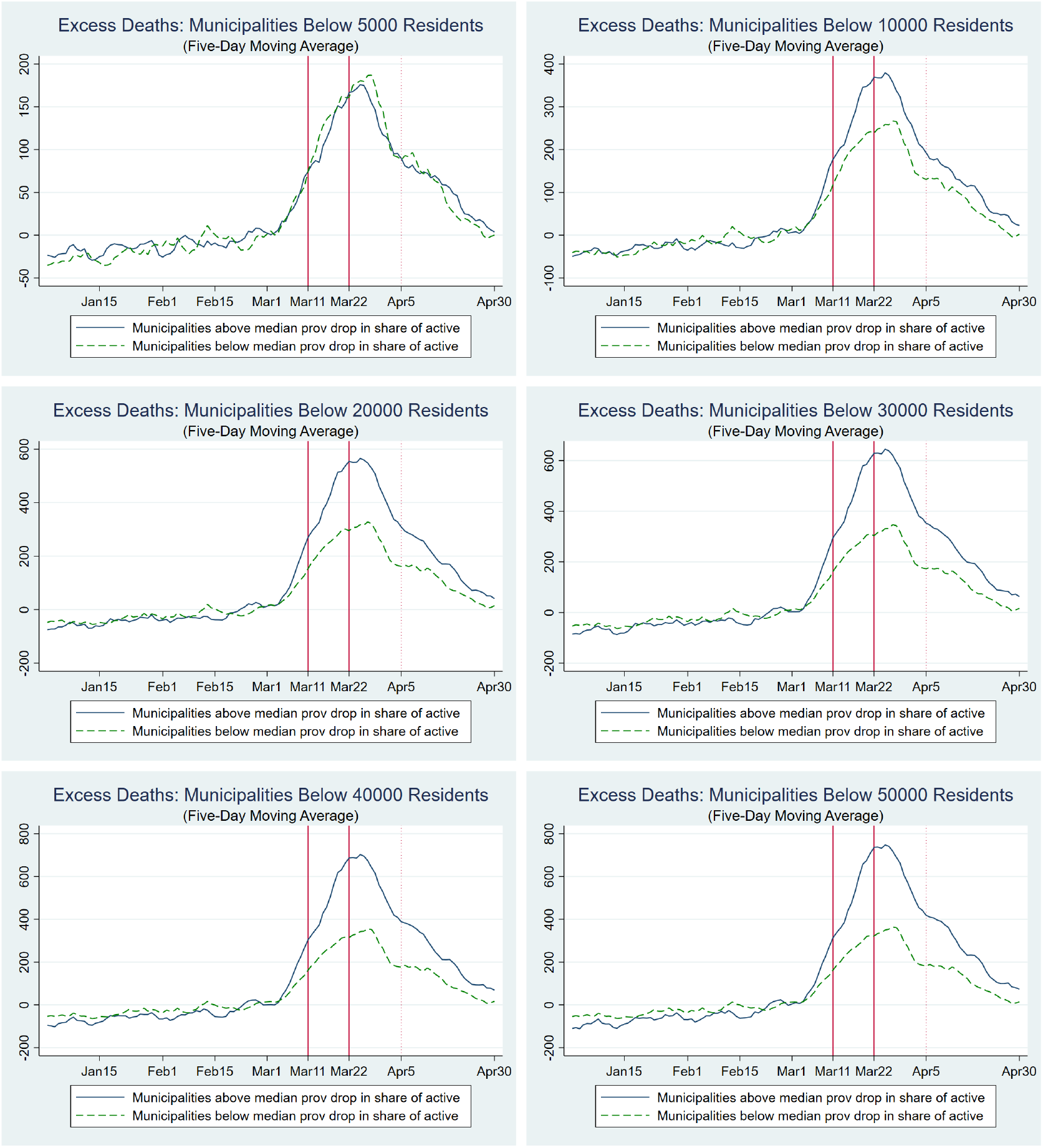
Excess Deaths: Subsamples by Municipal Population Threshold. Notes. The figure illustrates the evolution in the 5-day moving average of excess deaths at the municipal level in the treated group (blue line) and control group (green-dashed line) in different subsample below a given population threshold. The treated (control) group contains the municipalities above (below) the median drop in the share of active population in their province between the first and second lockdowns. The two vertical lines indicate the dates of the first (March 11) and second (March 22, economic) lockdown. The dotted vertical line indicates the end of the two-week gap after the economic lockdown (April 5).

The second alternative explanation is related to the effect of the first lockdown on mobility. Mobility is an important indicator of social interactions which, in turn, are key drivers of mortality by Covid-19 (Fang *et al*., 2020; Glaeser *et al*., 2020; Kraemer *et al*., 2020). As illustrated by Figure 2, before the first lockdown above-the-median municipalities had a consistently higher level of mobility (possibly because these municipalities had a higher share of active population absent lockdown measures, as also discussed in Weill *et al*. (2020) for the U.S.). At the same time, in the first lockdown period — following the shutdown of schools, restaurants, gyms, and other activities — the two groups converged in terms of mobility. This suggests that above-the-median municipalities experienced a stronger reduction in mobility already in the first lockdown. Indeed, as Cronin and Evans (2020) documented, a reduction in mobility takes place before the legal lockdown in response to the diffusion of the virus. Accordingly, one might argue that our estimates are due to a behavioral response in above-the-median municipalities, which would have led to a reduction in Covid-19 mortality even absent any formal lockdown. Three pieces of evidence are not consistent with this argument. First, as illustrated by Table 4, in a control period in which the lagged effects of the first-lockdown are likely to be already in place (we drop the first two weeks of the first-lockdown, March 11 and March 25 as in column 4), our main estimates are still significant and the size of the coefficient is similar to that of our benchmark specification. For the first lockdown to be the driver of our effects, we should see no significant differences between above-the-median and below-the-median municipalities when comparing the (control) period March 26-April 4 with the (treatment) period April 5-April 30, as the first lockdown should already manifest its effects in terms of excess deaths in such control period. Second, also the placebo exercise shown in Table 4 (Column 5) does not support the hypothesis that our estimates are driven by a lagged effect of the first lockdown. If this was the case, we should observe a significant effect on excess deaths when comparing the first lockdown period with the two weeks after the beginning of the economic lockdown (when the second lockdown should not have much of an effect yet and, vice-versa, a lagged effect of the first lockdown should have an arguably significant effect on excess deaths). The coefficient of such placebo exercise is smaller than the one in our baseline specification and not significant. Last but not least, as discussed in Section 4, while the activities (e.g., schools and restaurants) that closed in the first lockdown period created a substantial reduction in mobility, their overall impact in terms of contraction in active population was rather limited and, importantly, rather homogenous across municipalities, compared with the second lockdown.

Overall, while the strong reduction in mobility observed in the first lockdown period is likely to have reduced the overall mortality by Covid-19 (Gatto *et al*., 2020), it does not seem to have had a differential impact in above-the-median vs. below the median municipalities (there are no significant differences in the trends of excess deaths between the groups of municipalities in response to the first lockdown). In turn, this suggests that our measure of aggregate mobility captures a fraction of the social interactions that are relevant to the excess mortality measure. For example, although the first lockdown and the associated closure of schools is likely to have had a strong impact on mobility, it may have reflected a reduction of social interactions mainly for age groups less likely to be a source of Covid-19 contagion (Lee and Raszka, 2020; Ludvigsson, 2020; Wu *et al*., 2020) and also less affected by Covid-19 mortality (Dowd *et al*., 2020). In contrast, the second lockdown, and the associated reduction in mobility following the closing of all non-essential economic activities, is likely to have mainly reflected a reduction of social interactions among workers and age groups relatively more affected by Covid-19. Hence, aggregate changes in mobility may have different effects on Covid-19 contagion and mortality depending on the types of social interactions they imply (see also Glaeser *et al*. (2020) for evidence on the heterogeneity of the impact of mobility on Covid-19 contagion both across space and over time).13 To sum up, while we cannot clearly pin-down the exact mechanism, our main results on excess deaths are unlikely to be driven by the containment effects of the first lockdown.

## 6 Conclusions

In response to the Covid-19 outbreak, most countries imposed restrictions on the movements of people and lockdowns of economic activities. In order to evaluate the effectiveness of these measures a key challenge is the measurement of their intensity. In this paper we focus on Italy, one of the first country hit by the pandemic, because it offers an ideal setting to identify the causal effect of the lockdown measures on mortality by Covid-19 (and, implicitly, on contagion). Specifically, we exploit the exogenous variation of inactive population across Italian municipalities due to the selective and progressive restriction of sectors subject to the lockdown to measure its intensity.

Our main finding is to provide an estimate of the lives saved by the tightening of the economic lockdown at the peak of the pandemic in Italy. Consistently with the evidence that Covid-19 is particularly risky for older-age groups, we observe a statistically significant effect for the entire population and for age groups between 40-64 and older (with larger and more significant effects for the cohort above 50). In addition, we find that the effectiveness of the lockdown is related to the induced reduction in people mobility: municipalities with a larger contraction of the share of active population experienced a reduction in daily mobility of 53 kilometers per 1,000 residents with respect to municipalities with a smaller contraction. A simple back of the envelope calculation indicates that the intensity of the lockdown, proxied by the reduction in the share of active population by 42.5 instead of 17 percentage points, avoided 4,793 deaths in 3,518 municipalities between April 5 and April 30, or 1.36 lives per municipality. A one percentage point reduction in the share of active population translated into a 1.32 percentage points reduction in mortality by Covid-19.

Our results are informative of the cost-effectiveness of lockdown measures during the first wave of the pandemic. However, the results cannot be conclusive on whether or not the severe economic lockdown has been a desirable policy on the basis of a welfare analysis. On one hand, to assess the benefit of the lockdown in terms of economic value of reduced deaths, one would need to have detailed information on the socio-demographic characteristics on the people that lost their life during the first pandemic wave. Unfortunately, these data are not publicly available in the Italian context. On the other hand, one would have to estimate not only the cost of the economic lockdown in terms of permanent loss of output (and thus have detailed data on the value added at the municipality-economic sector level) but one would also have to compute the lockdown cost in terms of psychological stress suffered by the population. Hence, our analysis can be seen as a first step to address the cost-effectiveness of the Italian lockdown. Finally it should be clear that we cannot claim the same effects in terms of avoided deaths would necessarily hold in different settings, for example when masks are available (Lyu and Wehby, 2020a) or a better contact-tracing system is in place. More generally, our empirical strategy provides a simple methodology for future research aiming to assess the impact of the economic lockdown in reducing Covid-19 deaths in other countries.

## Data Availability

All data are publicly available

## Online Appendix

### Appendix A

#### A1. National level lockdowns in Italy

The chronology of the nation-wide “nonpharmaceutical interventions” (NPIs) in Italy is the following:^14^

- Schools (and major sport events) lockdown: On March 4, 2020 the Government approved a Decree ordering the closing of schools, universities, and football stadiums in all regions. In addition, it introduced restrictions to the access of relatives and visitors to hospitals and nursing homes.
- First lockdown: On March 11, 2020 the Government approved a Decree, publicly referred to as the *Decreto #IoRestoACasa* (or, “I stay home”). With this Decree all business activities open to the public were closed (with the exception of, mostly, grocery stores). All large public gatherings were also prohibited.
- Second Lockdown (or Economic Lockdown): On March 22, the Ministries of Interior and Health approved an order that prohibited any movement of people from their homes, with the exception of specific and proven work or health related reasons. On the same day, the Government approved a Decree that ordered the closing of all non essential economic activities.

Some of these lockdown measures were then relaxed starting on May 4, 2020 (this is the start of the so called “phase 2” of the lockdown)^15^.

#### A2. Data on share of active/inactive population

To calculate the share of active population in each municipality in the first lockdown period (March 11-21) and second lockdown period (March 22-April 30) we combine two sources of data. First, we gathered data on the number of workers in each municipality at the ATECO (i.e., the Italian equivalent of the European NACE) 3-digit sectors. The latest data are available for 2017 through the “Registro Statistico delle Unita` Locali” (ASIA UL)^16^. As no major changes have occurred to the Italian economy between 2017 and the start of the pandemic in 2020, this dataset provides a reliable measure of the number of active workers in each municipality before the lockdowns. Then, to construct a variable capturing the share of active population in each municipality before the lockdowns, we simply divide the number of active workers by the municipal population. We gathered data on the ATECO sectors that were active/inactive during the first and second lockdown by looking at the list of active/inactive sectors specified in the Government Decrees which introduced the lockdown measures. Specifically, the March 11 and March 22 Decrees both specified detailed categories of ATECO sectors that were allowed/not allowed to operate^17^. We then simply combined these two sources of data and computed the share of active population in each municipality during the first and second lockdown^18^. Our key variable (the difference between the active population shares in the second lockdown compared with the first lockdown) is then computed by taking the variation in active population after the second lockdown.

### Appendix B. Excess Deaths

This section evaluates the robustness of our results in terms of the effect of the lockdown of economic activities on excess mortality, and then presents additional results.

#### B2. Additional Robustness Checks

Table B.1 presents results when considering the variation in the share of active workers rather than in the share of active population (i.e., considering only the total municipal labour force population as a denominator).

In addition, we investigate to what extent our findings are sensitive to the use of the median value of the drop in the share of active population (in the province) as a threshold to categorize our municipalities as *treated*. To do so, we consider two alternative specifications that look at two different thresholds: the 25th percentile, and the 75th percentile. Just like in the main analysis, our coefficient of interest will be *L*^25^ (*L*^75^), which now takes value 1 if the municipality has experienced a drop in the share of active workers above the 25th (75th) percentile in the province and if *t ≥ t*_0_+14. Results are reported in Table B.2, where we evaluate the effect of the so-defined *L* on total excess deaths. Consistently with the claim that municipalities that experienced a stronger reduction in active population saw a larger decrease in excess deaths (with respect to those municipalities who did not), the size of the coefficients show that the effect of the lockdown increases as we consider as treated those municipalities with more pronounced drops in active population.

Table B.3 provides a further age breakdown relative to our results on the impact of the economic lockdown on the excess mortality of the population aged 30-64 (as shown in Table 2 in the paper). The breakdown shows that, as expected, the economic lockdown had a more significant effect on the older cohorts.

The next two tables, instead, replicate the analysis in Table 2 distinguishing for gender, that is considering only males in Table B.4 and females in Table B.5. As we can see, across agegroups, the economic lockdown seems to have had similar effects on males and females, with the exception of the 30-64 cohort showing more significant effects for males.

Finally, Table B.6 provides further evidence that our results are robust to several additional specifications and extensions. Namely, Column 1 presents results when clustering at the labor market area (LMAs) level rather than at the (smaller) municipal level, thus accounting for potential spatial spillovers in commuting areas. In Column 2 the sample is limited to the municipalities for which we have complete daily data on mobility from EnelX; Column 3 provides results for the sample where we drop provincial capital municipalities; Column 4 presents results only for the municipalities belonging to the Lombardy region (i.e., the Italian region most affected by the Covid-19 shock), whereas Column 5 provides the results when excluding them; Column 6 reports results using the growth rate in excess mortality as the dependent variable.

#### B2. Robustness: Subsamples by percentiles drop in the share of active population

In this section we show to what extent our results are robust to narrowing our sample around the provincial median drop in the share of active population.

Table B.7 shows that our key results are, in fact, robust to such test. In particular, it is still possible to detect a negative and statistically significant effect of the economic lockdown on excess deaths even when looking at municipalities between the 30th and 70th percentile of the drop in the share of active in their province. That is, when considering municipalities with a drop above the median and below the 70th percentile as treated, and municipalities below the median and above the 30th percentile as control.

These results provide an important robustness check for our baseline estimates since, as illustrated in Figure 3, the parallel trend assumption seems more plausible in the subsample of municipalities in the 30th-70th percentiles range. We should also notice that results are not significant anymore when futher narrowing the sample to municipalities in the 40th-60th percentiles range. This is likely due to both the rather small number of municipalities in this sample (around 1,335) and to the limited difference in the drop in the share of active between above and below-the-median municipalities.

**Table B.1:**
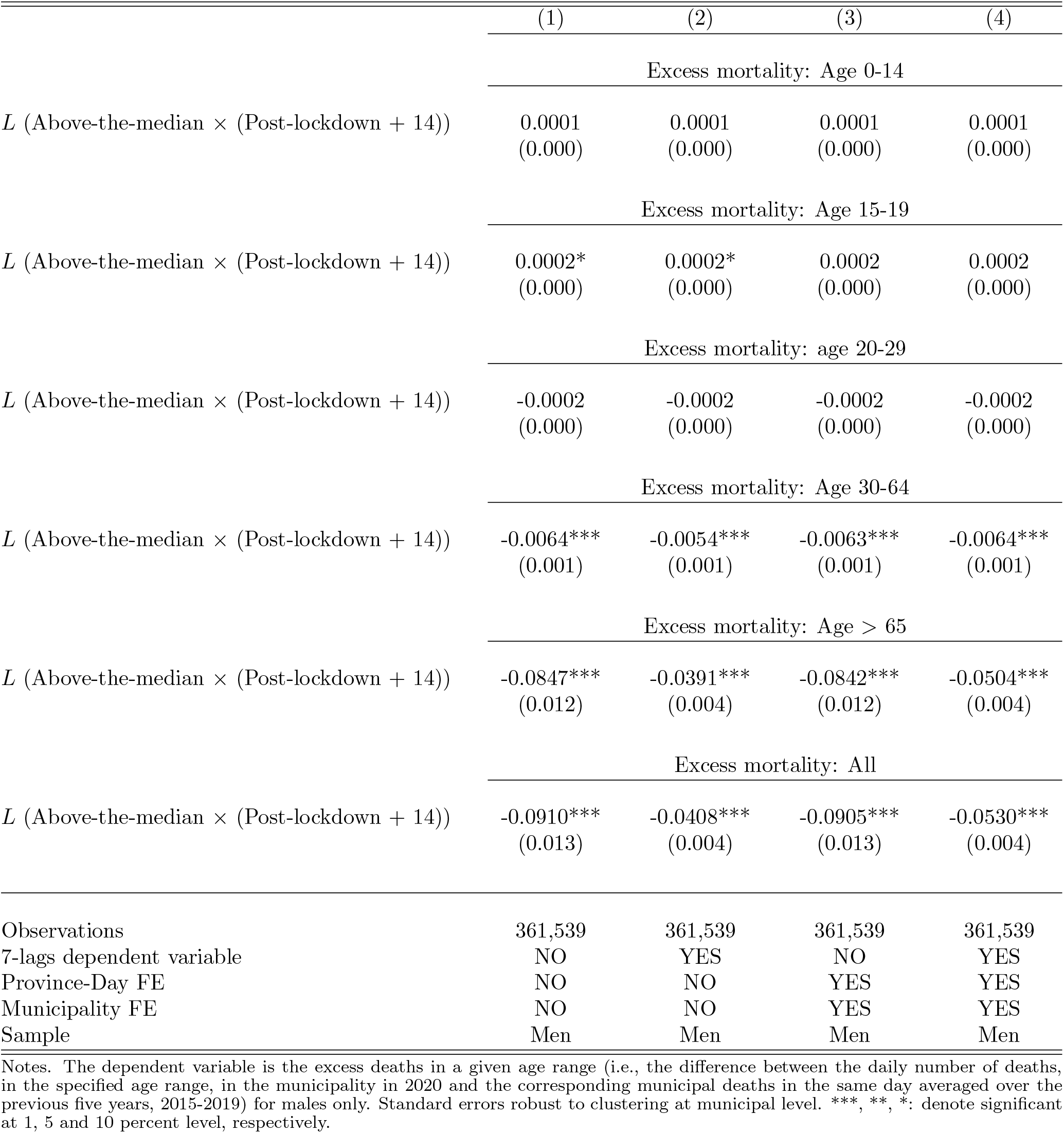
Economic Lockdown and Excess Deaths - Variation in share of active workers

#### B3. Robustness: Subsamples by municipal population

Table B.8 shows that our key results are robust to restricting the analysis to these different samples with respect to municipal population. These results provide a further key robustness check of our baseline estimates since, as illustrated in Figure 4, the parallel trend assumption seems to be satisfied in the subsamples of municipalities below smaller population thresholds.

**Table B.2:** Economic Lockdown and Excess Deaths - Alterntive thresholds

|  | (1) | (2) | (3) | (4) |
| --- | --- | --- | --- | --- |
|  | Above vs Below 25th percentile |  |  |  |
| $L^{25}$ (Above-the-25th pct × (Post-lockdown + 14)) | -0.0808***<br>(0.009) | -0.0325***<br>(0.003) | -0.0789***<br>(0.009) | -0.0439***<br>(0.004) |
|  | Above vs Below 75th percentile |  |  |  |
| $L^{75}$ (Above-the-75th pct × (Post-lockdown + 14)) | -0.1187***<br>(0.024) | -0.0510***<br>(0.008) | -0.1191***<br>(0.023) | -0.0684***<br>(0.009) |
| Observations | 361,539 | 361,539 | 361,539 | 361,539 |
| 7-lags dependent variable | NO | YES | NO | YES |
| Province-Day FE | NO | NO | YES | YES |
| Municipality FE | NO | NO | YES | YES |

Notes. The dependent variable is the total excess deaths (i.e., the difference between the daily number of total deaths in the municipality in 2020 and the corresponding municipal deaths in the same day averaged over the previous five years, 2015-2019). Standard errors robust to clustering at municipality level. \*\*\*, \*\*, \*: denote significant at 1, 5 and 10 percent level, respectively.

#### B4. Robustness: Geographical Heterogeneity

Table B.9 shows the heterogeneity of our results in different Italian macro-regions (North, Center and South). The results show that the economic lockdown was effective in reducing Covid-19 mortality in the North and in the Center. The estimates for the South are also negative, yet not statistically significant. This may easily be explained by the fact that, at the time of the lockdown, the diffusion of the epidemic was very limited in the South. At the same time, the absence of a significant effect in the South tends to exclude other confounding mechanisms unrelated to Covid-19 (e.g., a reduction of deaths due to a reduction of traffic accidents and/or to a reduction of workplace accidents).

**Table B.3:** Economic Lockdown and Excess Deaths (Adults)

|  | (1) | (2) | (3) | (4) |
| --- | --- | --- | --- | --- |
| Excess mortality: Age 30-39 |  |  |  |  |
| $L$ (Above-the-median × (Post-lockdown + 14)) | -0.0003<br>(0.000) | -0.0003<br>(0.000) | -0.0003<br>(0.000) | -0.0004<br>(0.000) |
| Excess mortality: Age 40-49 |  |  |  |  |
| $L$ (Above-the-median × (Post-lockdown + 14)) | -0.0009*<br>(0.000) | -0.0009**<br>(0.000) | -0.0009*<br>(0.000) | -0.0009*<br>(0.001) |
| Excess mortality: Age 50-64 |  |  |  |  |
| $L$ (Above-the-median × (Post-lockdown + 14)) | -0.0051***<br>(0.001) | -0.0044***<br>(0.001) | -0.0050***<br>(0.001) | -0.0052***<br>(0.001) |
| Observations | 361,539 | 361,539 | 361,539 | 361,539 |
| 7-lags dependent variable | NO | YES | NO | YES |
| Province-Day FE | NO | NO | YES | YES |
| Municipality FE | NO | NO | YES | YES |

Notes. The dependent variable is the excess deaths in a given age range (i.e., the difference between the daily number of deaths, in the specified age range, in each municipality in 2020 and the corresponding municipal deaths in the same day averaged over the previous five years, 2015-2019). Standard errors robust to clustering at municipality level. \*\*\*, \*\*, \*: denote significant at 1, 5 and 10 percent level, respectively.

**Table B.4:** Economic Lockdown and Excess Deaths - Male population

|  | (1) | (2) | (3) | (4) |
| --- | --- | --- | --- | --- |
| Excess mortality: Age 0-14 |  |  |  |  |
| $L$ (Above-the-median × (Post-lockdown + 14)) | 0.0001<br>(0.000) | 0.0001<br>(0.000) | 0.0001<br>(0.000) | 0.0001<br>(0.000) |
| Excess mortality: Age 15-19 |  |  |  |  |
| $L$ (Above-the-median × (Post-lockdown + 14)) | 0.0001*<br>(0.000) | 0.0001*<br>(0.000) | 0.0001*<br>(0.000) | 0.0002*<br>(0.000) |
| Excess mortality: age 20-29 |  |  |  |  |
| $L$ (Above-the-median × (Post-lockdown + 14)) | -0.0002*<br>(0.000) | -0.0002*<br>(0.000) | -0.0002*<br>(0.000) | -0.0003*<br>(0.000) |
| Excess mortality: Age 30-64 |  |  |  |  |
| $L$ (Above-the-median × (Post-lockdown + 14)) | -0.0051***<br>(0.001) | -0.0046***<br>(0.001) | -0.0050***<br>(0.001) | -0.0052***<br>(0.001) |
| Excess mortality: Age > 65 |  |  |  |  |
| $L$ (Above-the-median × (Post-lockdown + 14)) | -0.0562***<br>(0.008) | -0.0234***<br>(0.002) | -0.0561***<br>(0.007) | -0.0345***<br>(0.003) |
| Excess mortality: All |  |  |  |  |
| $L$ (Above-the-median × (Post-lockdown + 14)) | -0.0612***<br>(0.008) | -0.0256***<br>(0.002) | -0.0610***<br>(0.008) | -0.0371***<br>(0.003) |
| Observations | 361,539 | 361,539 | 361,539 | 361,539 |
| 7-lags dependent variable | NO | YES | NO | YES |
| Province-Day FE | NO | NO | YES | YES |
| Municipality FE | NO | NO | YES | YES |
| Sample | Men | Men | Men | Men |

Notes. The dependent variable is the excess deaths in a given age range (i.e., the difference between the daily number of deaths, in the specified age range, in the municipality in 2020 and the corresponding municipal deaths in the same day averaged over the previous five years, 2015-2019) for males only. Standard errors robust to clustering at municipal level. \*\*\*, \*\*, \*: denote significant at 1, 5 and 10 percent level, respectively.

### Appendix C. Mobility

This section presents evidence that the results in the paper are robust to an alternative measure of mobility. In addition, we assess the reliability of the mobility data using an alternative data source for a subset of the municipalities in our sample.

**Table B.5:**
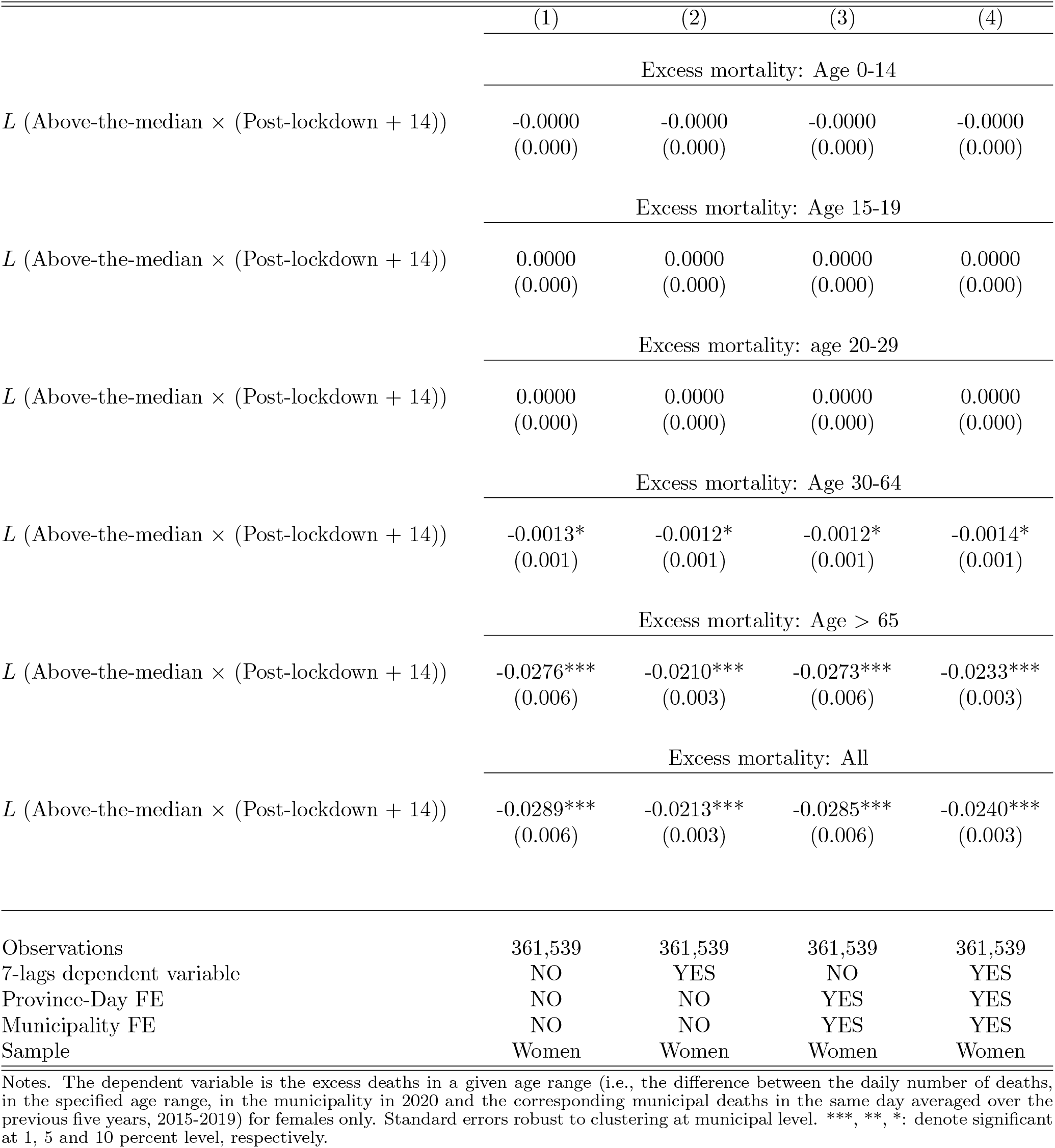
Economic Lockdown and Excess Deaths - Female population

**Table B.6:** Economic Lockdown and Excess Deaths: Additional Robustness

|  | (1)<br>Cluster<br>LMAs | (2)<br>Mobility<br>Sample | (3)<br>Drop<br>Provincial<br>Capitals | (4)<br>Only<br>Lombardy | (5)<br>Drop<br>Lombardy | (6)<br>Growth<br>Rate |
| --- | --- | --- | --- | --- | --- | --- |
| $L$ (Above-the-median<br>× (Post-lockdown + 14)) | -0.0530***<br>(0.005) | -0.0565***<br>(0.006) | -0.0432***<br>(0.005) | -0.0995***<br>(0.011) | -0.0363***<br>(0.005) | -0.0115***<br>(0.002) |
| Observations | 358,377 | 273,309 | 353,073 | 73,542 | 287,895 | 334,334 |
| 7-lags dependent variable | YES | YES | YES | YES | YES | YES |
| Province-Day FE | YES | YES | YES | YES | YES | YES |
| Municipality FE | YES | YES | YES | YES | YES | YES |

Notes. In Columns 1-5, the dependent variable is the total excess deaths (i.e., the difference between the daily number of deaths in each municipality in 2020 and the total municipal deaths in the same day averaged over the previous five years, 2015-2019). In Column 6, the dependent variable is the daily municipal growth rate in total excess mortality. Column 1 presents results when clustering at the labor market areas (LMAs) level rather than at the municipal level. Column 2 provides results for the sample of municipalities with complete mobility data. In Column 3 the sample excludes provincial capitals. Column 4 presents results only for the municipalities belonging to the Lombardy region, whereas Column 5 provides the results when excluding them. Standard errors robust to clustering at municipality level in Columns 2-6. \*\*\*, \*\*, \*: denote significant at 1, 5 and 10 percent level, respectively.

#### C1. Robustness

In the paper we use a measure of mobility based on the number of kilometers divided by population obtained from EnelX. In this section we show that this measure is very similar to an alternative measure based on the number of “movements”, divided by population, also from EnelX. Specifically, Figure C.1 plots the time variation in mobility – in municipalities above/below the provincial median reduction in the share of active population – using the number of movements per 1,000 residents.^19^ It is easy to see that the pattern is very close to the one illustrated in Figure 2 in the paper when taking the kilometers per 1,000 residents.

Similarly, Table C.1 presents evidence on the effect of the economic lockdown on the number of movements per 1,000 residents during the weekdays (Monday-Friday) and in the weekends (Saturday-Sunday). Again, it is easy to see that results are very similar to those presented in Table 1 in the paper when taking the kilometers per 1,000 residents as a mobility indicator. Specifically, the economic lockdown has a negative and significant effect on the number of movements, and especially so in the weekdays.

**Table B.7:**
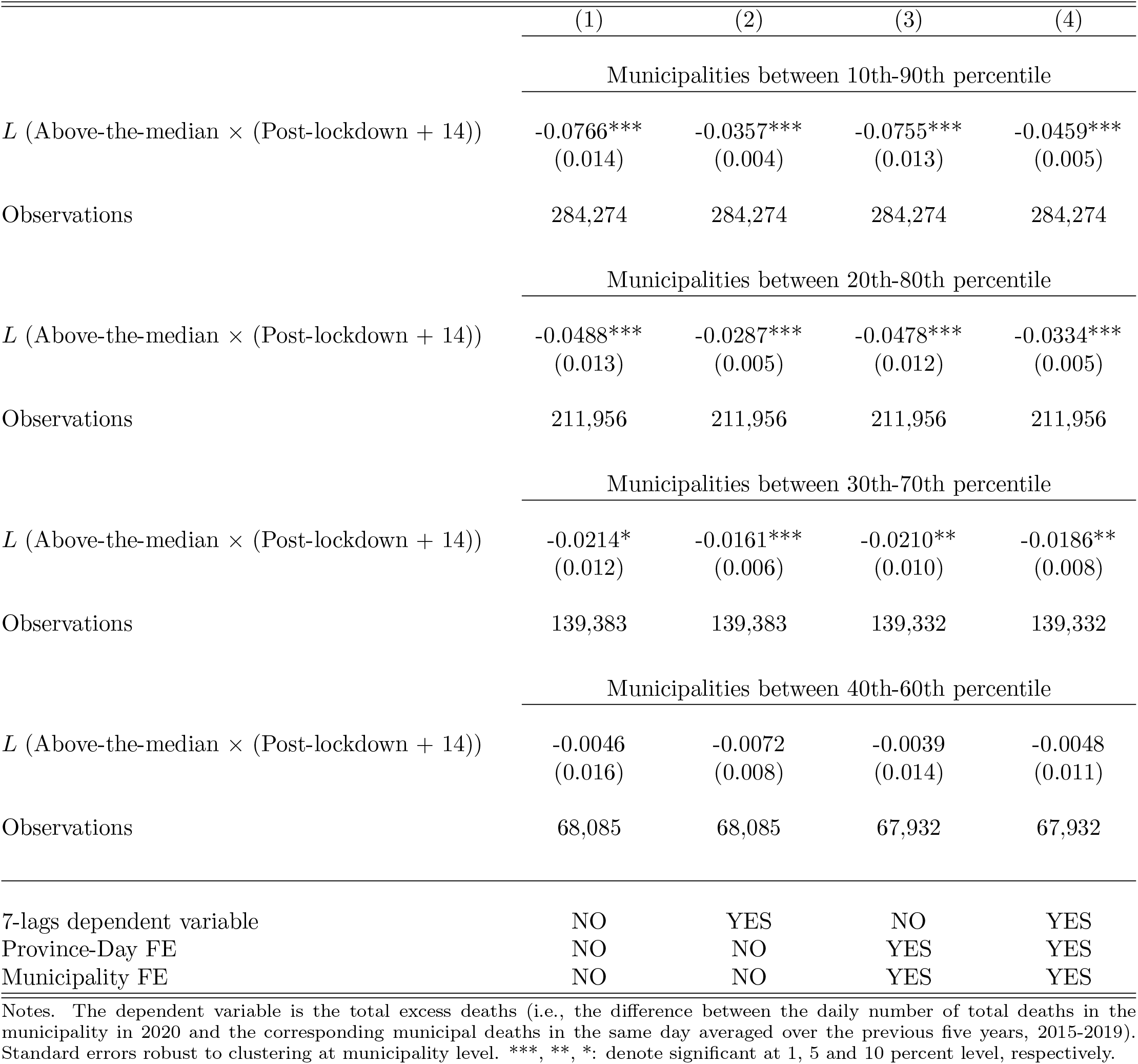
Economic Lockdown and Excess Deaths - Subsamples by percentile drop share of active in the province

**Table B.8:** Economic Lockdown and Excess Deaths - Subsamples by Municipal Population

|  | (1) | (2) | (3) | (4) |
| --- | --- | --- | --- | --- |
| Municipalities below 5000 residents |  |  |  |  |
| $L$ (Above-the-median × (Post-lockdown + 14)) | -0.0085**<br>(0.004) | -0.0067**<br>(0.003) | -0.0091**<br>(0.004) | -0.0089**<br>(0.004) |
| Observations | 252,603 | 252,603 | 252,501 | 252,501 |
| Municipalities below 10000 residents |  |  |  |  |
| $L$ (Above-the-median × (Post-lockdown + 14)) | -0.0291***<br>(0.006) | -0.0180***<br>(0.003) | -0.0264***<br>(0.005) | -0.0237***<br>(0.004) |
| Observations | 305,388 | 305,388 | 305,337 | 305,337 |
| Municipalities below 20000 residents |  |  |  |  |
| $L$ (Above-the-median × (Post-lockdown + 14)) | -0.0438***<br>(0.007) | -0.0257***<br>(0.003) | -0.0416***<br>(0.006) | -0.0351***<br>(0.004) |
| Observations | 337,212 | 337,212 | 337,212 | 337,212 |
| Municipalities below 30000 residents |  |  |  |  |
| $L$ (Above-the-median × (Post-lockdown + 14)) | -0.0464***<br>(0.007) | -0.0270***<br>(0.003) | -0.0447***<br>(0.006) | -0.0375***<br>(0.005) |
| Observations | 346,902 | 346,902 | 346,902 | 346,902 |
| Municipalities below 40000 residents |  |  |  |  |
| $L$ (Above-the-median × (Post-lockdown + 14)) | -0.0495***<br>(0.007) | -0.0287***<br>(0.003) | -0.0484***<br>(0.006) | -0.0402***<br>(0.005) |
| Observations | 351,900 | 351,900 | 351,900 | 351,900 |
| Municipalities below 50000 residents |  |  |  |  |
| $L$ (Above-the-median × (Post-lockdown + 14)) | -0.0523***<br>(0.007) | -0.0307***<br>(0.003) | -0.0514***<br>(0.006) | -0.0426***<br>(0.005) |
| Observations | 354,552 | 354,552 | 354,552 | 354,552 |
| 7-lags dependent variable | NO | YES | NO | YES |
| Province-Day FE | NO | NO | YES | YES |
| Municipality FE | NO | NO | YES | YES |

Notes. The dependent variable is the total excess deaths (i.e., the difference between the daily number of total deaths in the municipality in 2020 and the corresponding municipal deaths in the same day averaged over the previous five years, 2015-2019). Standard errors robust to clustering at municipality level. \*\*\*, \*\*, \*: denote significant at 1, 5 and 10 percent level, respectively.

**Table B.9:**
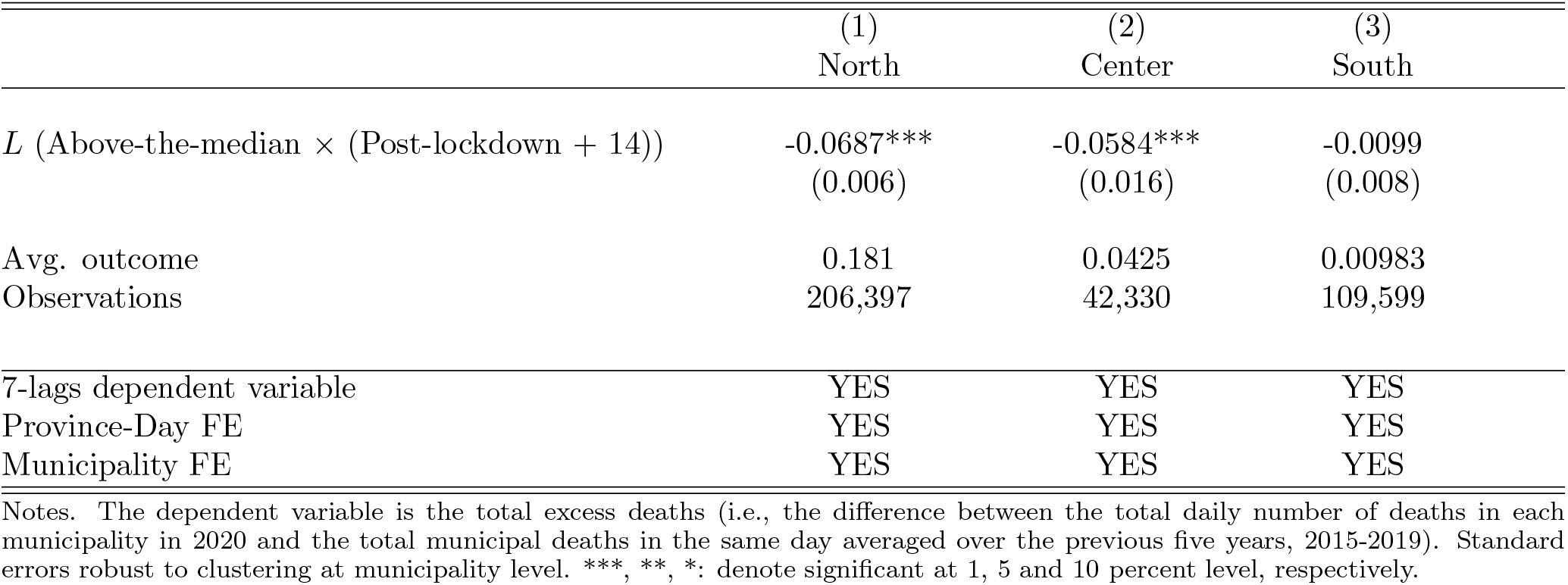
Economic Lockdown and Excess Deaths by Macro-Region

|  | (1)<br>North | (2)<br>Center | (3)<br>South |
| --- | --- | --- | --- |
| $L$ (Above-the-median × (Post-lockdown + 14)) | -0.0687***<br>(0.006) | -0.0584***<br>(0.016) | -0.0099<br>(0.008) |
| Avg. outcome | 0.181 | 0.0425 | 0.00983 |
| Observations | 206,397 | 42,330 | 109,599 |
| 7-lags dependent variable | YES | YES | YES |
| Province-Day FE | YES | YES | YES |
| Municipality FE | YES | YES | YES |

Notes. The dependent variable is the total excess deaths (i.e., the difference between the total daily number of deaths in each municipality in 2020 and the total municipal deaths in the same day averaged over the previous five years, 2015-2019). Standard errors robust to clustering at municipality level. \*\*\*, \*\*, \*: denote significant at 1, 5 and 10 percent level, respectively.

**Figure C.1:**
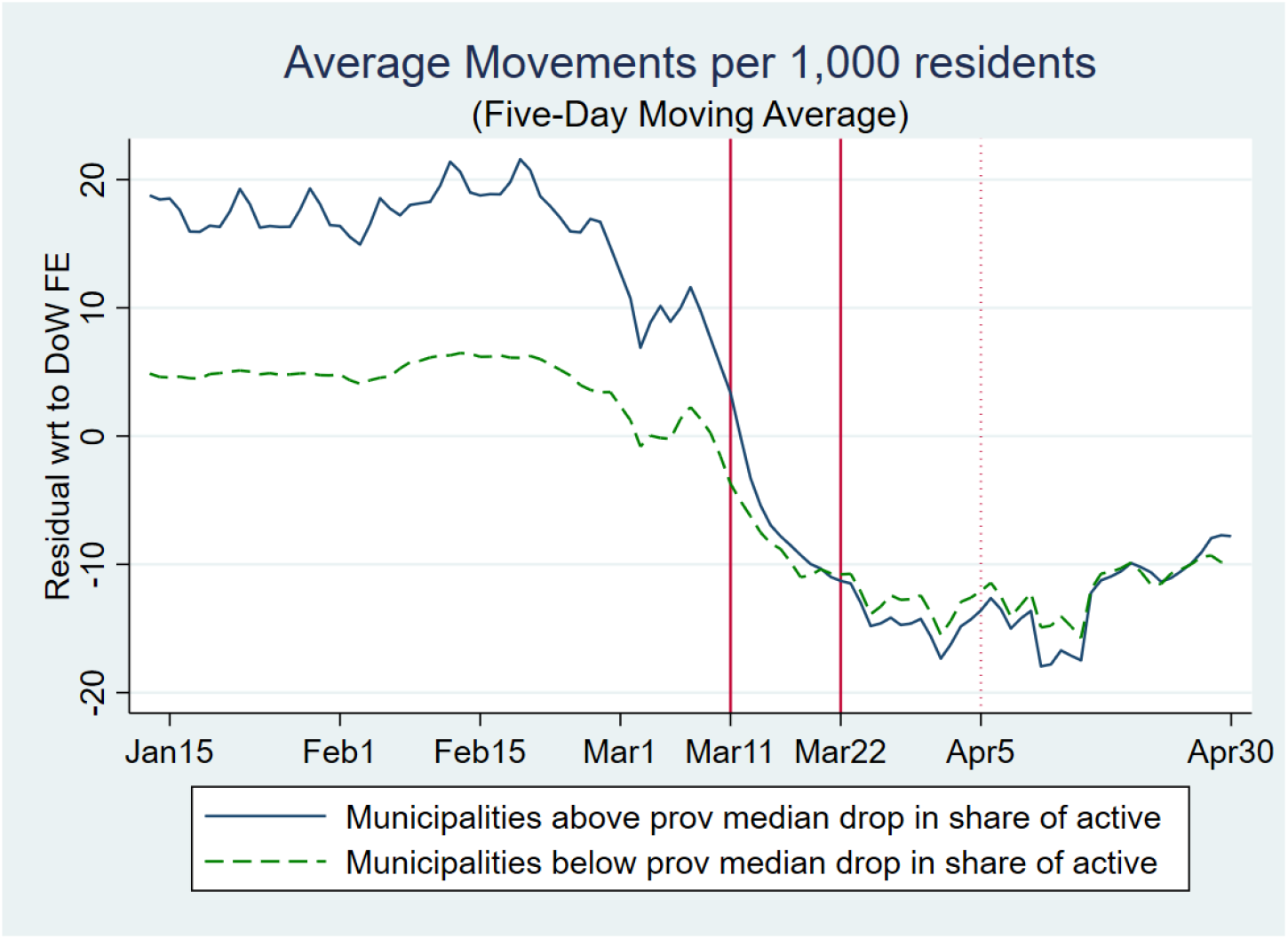
Movements per capita. Notes. The figure illustrates the time-variation in the 5-day moving average in the movements per 1,000 residents (as residuals with respect to day-of-the-week fixed effects) at the municipal level in the treated group (solid blue line) and control group (green dashed line). The treated (control) group contains the municipalities above (below) the median with respect to the drop in the share of active population in their province between the first and second lockdowns. The two solid vertical lines indicate the dates of the first and second (or economic) lockdown. The mobility data are from EnelX.

#### C2. Validation

In this section we provide evidence that validates the reliability of the mobility data used in this paper, obtained from EnelX. Specifically, we show that various indicators of mobility, for a subset of the municipalities in our sample, are highly correlated to those constructed from an alternative data source. In particular, we obtain from Facebook, through its Data For Good program and Disease Prevention Maps, information on the number of users moving in and out of a given municipality for a given day.^20^ Unfortunately, the Facebook data has a smaller coverage in terms of municipalities with respect of the EnelX data. In fact, because of privacy concerns, Facebook does not report data when the number of individual users in smaller than 10. For this reason, in order to assess the quality of the baseline mobility data used in our paper, we decided to compare mobility indicators built with the two alternative data sources only for the larger municipalities. Figure C.2 and C.3 provide such comparison for the EnelX kilometers per 1,000 residents and movements per 1,000 residents, respectively, with the Facebook movements (in & out of a municipality) per 1,000 residents for a selected sample of such large municipalities (Milano, Roma, Firenze, Torino, Napoli and Venezia). For both measures, the scatter plots indicate a positive correlation for all municipalities.

**Table C.1:** Economic Lockdown and Mobility (movements per 1,000 residents)

|  | (1) | (2) |
| --- | --- | --- |
|  | Monday-Friday |  |
| $L$ (Above-the-median × Post-lockdown) | -3.7350***<br>(0.691) | -4.0673***<br>(0.680) |
| Avg. outcome | 48.76 | 48.76 |
| Observations | 198,236 | 198,236 |
|  | Saturday-Sunday |  |
| $L$ (Above-the-median × Post-lockdown) | -0.5224*<br>(0.280) | -0.5993**<br>(0.257) |
| Avg. outcome | 13.35 | 13.35 |
| Observations | 74,750 | 74,750 |
| Province-Day FE | NO | YES |
| Municipality FE | NO | YES |

Notes. The dependent variable is the number of movements per 1,000 residents. Standard errors robust to clustering at municipality level. \*\*\*, \*\*, \*: denote significant at 1, 5 and 10 percent level, respectively. Data are from EnelX-Here Technologies.

**Figure C.2:**
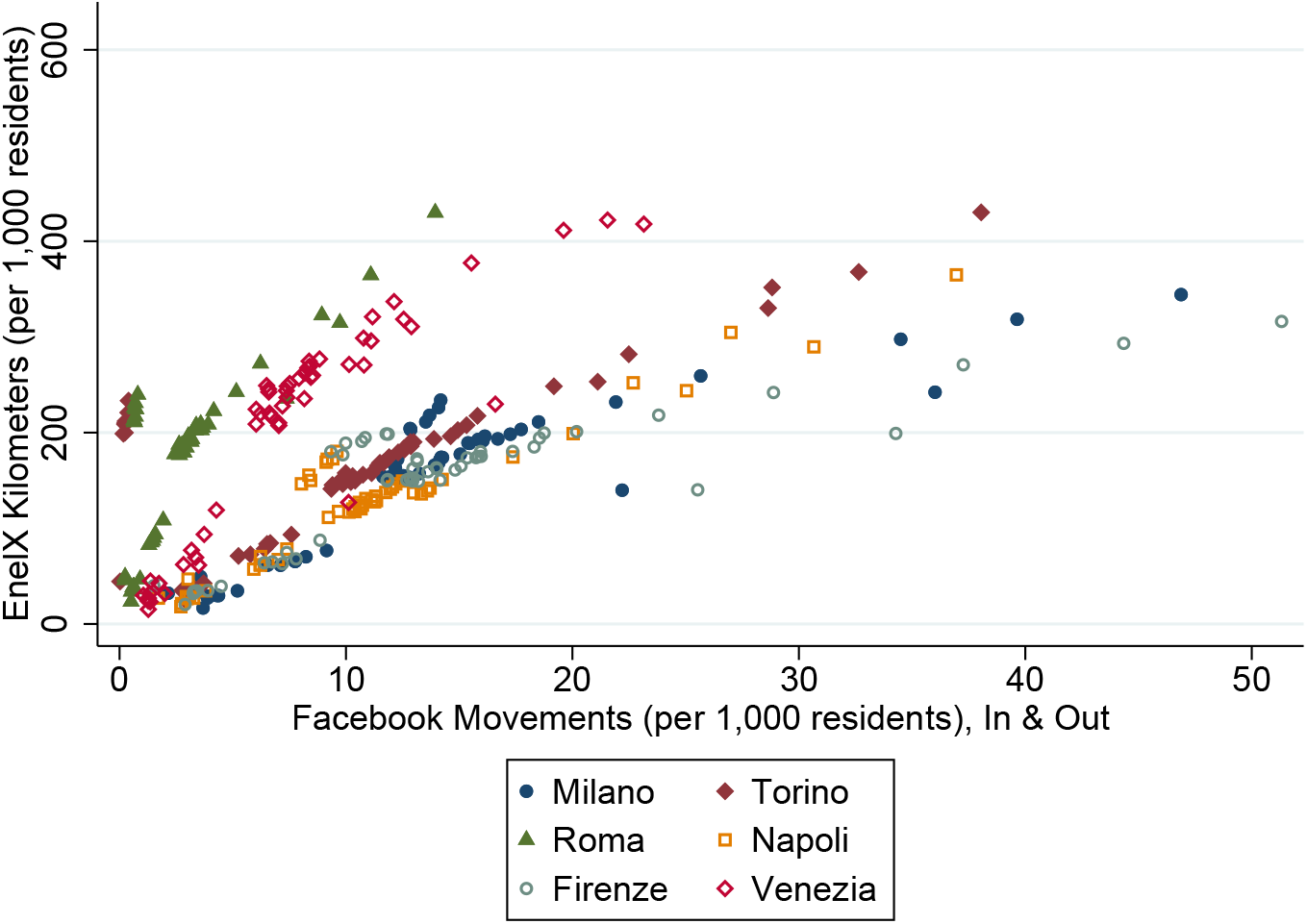
Kilometers (EnelX) and Movements in & out (Facebook) Notes. This figure plots the kilometers per 1,000 residents from EnelX against the mobility in or out of the municipality (also per 1,000 residents) constructed using the Facebook data.

**Figure C.3:**
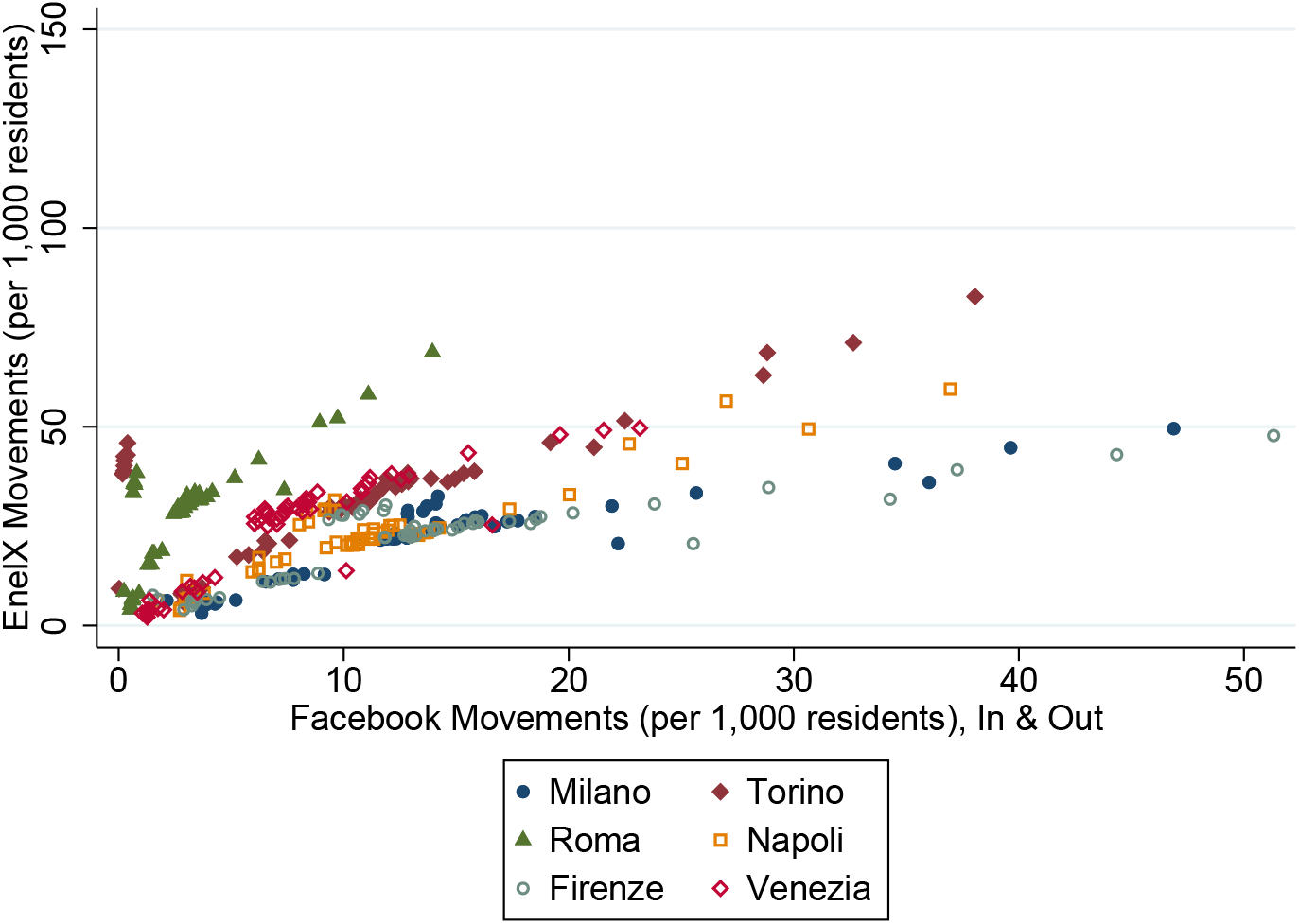
Movements (EnelX) and Movements in & out (Facebook) Notes. This figure plots the movements per 1,000 residents from EnelX against the mobility in or out of the municipality (also per 1,000 residents) constructed using the Facebook data.

Facebook also reports the percentage of users “who stay put” within a single location (in the Movement Range Maps). We build a daily indicator, at the province level, of Facebook users who move (1-% of users who stay put) and then compare it to the two indicators built with data from EnelX (i.e., kilometers or movements per 1,000 residents).^21^ Figures C.4 and C.5 provide additional evidence on the positive correlation between the mobility measures built with data from the two alternative sources EnelX mobility indicators with the one provided by Facebook.^22^ Finally, Table C.2 presents additional evidence that a positive and statistically significant correlation exists between these mobility indicators even when including province and date fixed effects.

**Figure C.4:**
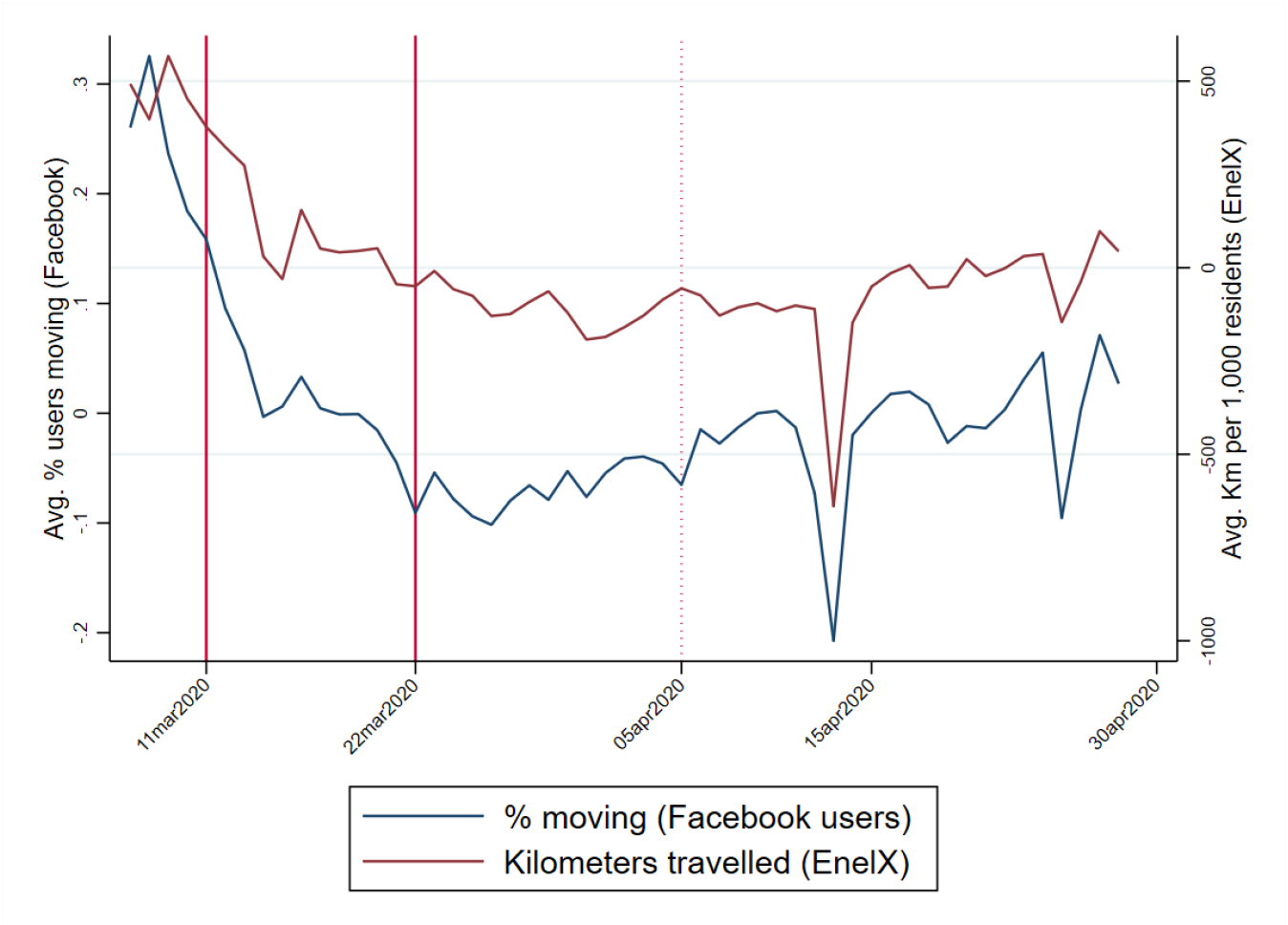
Km per 1,000 (EnelX) and % of Users moving (Facebook) Notes. This figure plots the kilometers per 1,000 residents from EnelX and (solid red line) and the percentage of Facebook users that are moving (solid blue line). The two solid vertical lines indicate the dates of the first and second (or economic) lockdown. Data are from EnelX-Here Technologies and Facebook.

**Figure C.5:**
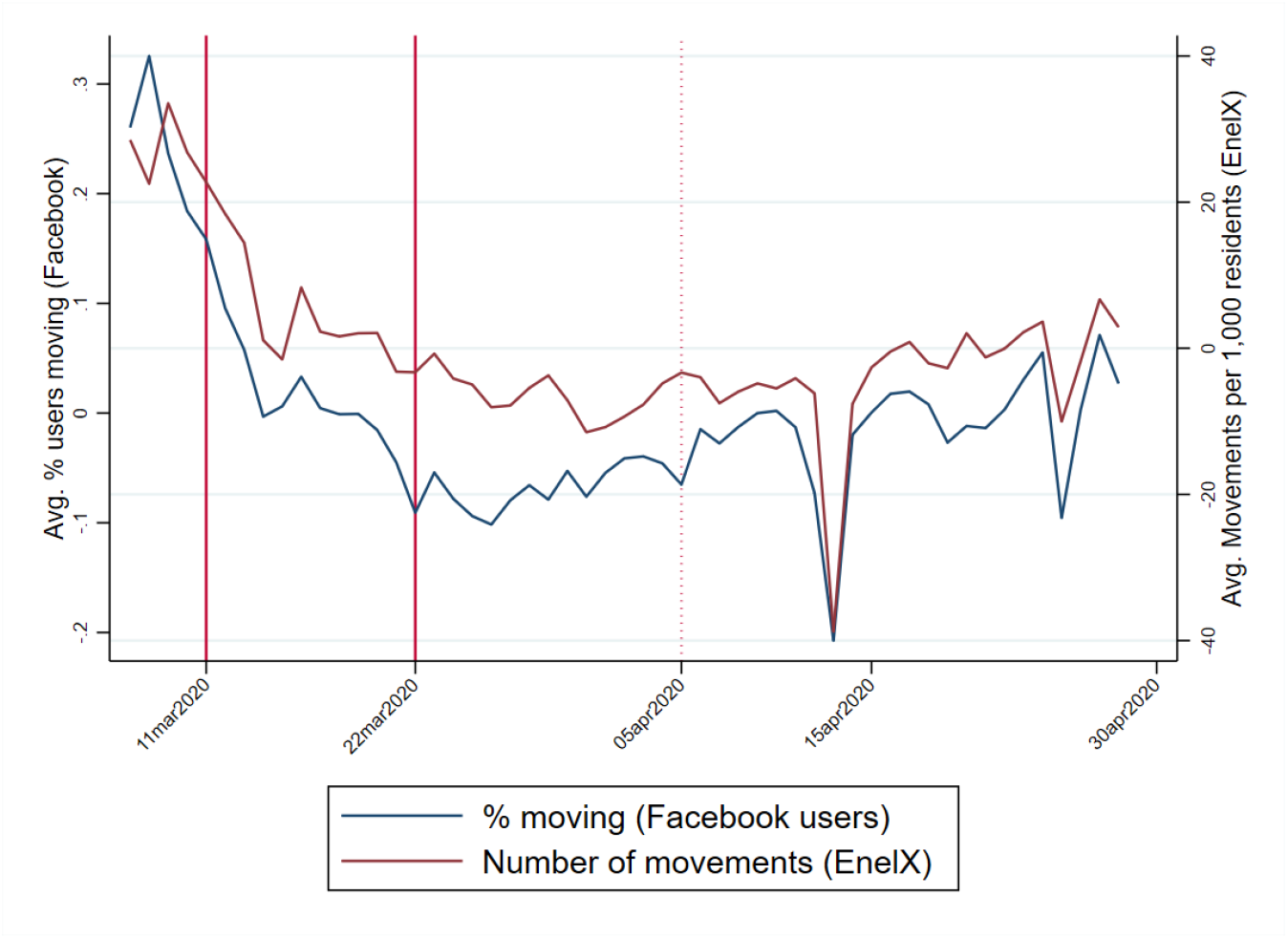
Movements per 1,000 (EnelX) and % of Users moving (Facebook) Notes. This figure plots the movements per 1,000 residents from EnelX (solid red line) and the percentage of Facebook users that are moving (solid blue line). The two solid vertical lines indicate the dates of the first and second (or economic) lockdown. Data are from EnelX-Here Technologies and Facebook.

**Table C.2:** EnelX vs. Facebook Mobility Data

|  | (1) | (2) | (3) | (4) | (5) | (6) |
| --- | --- | --- | --- | --- | --- | --- |
|  | EnelX |  |  | EnelX |  |  |
|  | Km per 1,000 |  |  | Movements per 1,000 |  |  |
| Facebook | 3,253.9*** | 3,084.2*** | 3,273.6*** | 187.4*** | 188.0*** | 172.5*** |
| % moving users | (88.979) | (254.590) | (683.180) | (4.003) | (10.054) | (27.889) |
| Observations | 5,124 | 5,124 | 5,124 | 5,124 | 5,124 | 5,124 |
| Province FE | NO | YES | YES | NO | YES | YES |
| Date FE | NO | NO | YES | NO | NO | YES |
| Cluster at Province Level | NO | YES | YES | NO | YES | YES |

Notes. The dependent variable is number of kilometers (Columns 1-3) or Movements (Columns 4-6) per 1,000 residents in the province. \*\*\*, \*\*, \*: denote significant at 1, 5 and 10 percent level, respectively. Data are from EnelX-Here Technologies and Facebook.

### Appendix D. Fixed Effects and Lagged Excess Mortality

The model discussed in the paper includes municipality, province-by-date fixed effects and lagged values of the dependent variable (see Equation 1). As we mention in the paper, this general formulation has a demanding identifying assumption of sequential exogeneity. However, variations of Model 1 that include either fixed effects or lagged values of *ED* have a bracketing property that provide bounds to the causal effect of *L*_*it*_ on excess mortality (*β*). In this section we closely follow Angrist and Pischke (2008) and Guryan (2001) to explain this bracketing property.

To simplify the exposition we do not consider province-by-date fixed effects (thus we ignore the *j*-province dimension) and we consider only two periods, a pre-treatment one where *L* is equal to zero for all municipalities and a post-treatment in which *L* is equal to 1 only for the above-the-median municipalities. The results can be generalized to *n* periods. We start the discussion assuming that the excess number of deaths *ED* is described according to the following equations:

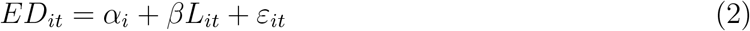

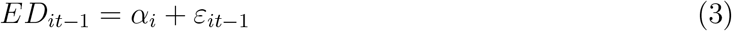

where *L*_*it*_ is correlated with municipality fixed effects *α*_*i*_. In this formulation differences in *ED* between above and below-the-median municipalities in the pre-treatment period depend on fixed characteristics. In the second period, further differences in *ED* are captured by the estimated *β*. In addition, consider *ε*_*it*_ to be uncorrelated with both *α*_*i*_ and *L*_*it*_.

Let’s assume that, although the true model is described by 2 and 3, we fail to estimate the right model and instead estimate a model with lagged *ED* but no municipality fixed effects:

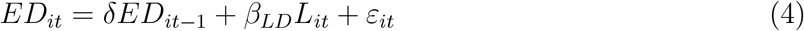

In this case, exploiting 2 and 3 and re-arranging we have an estimated *β*_*LD*_ from 4 such that:

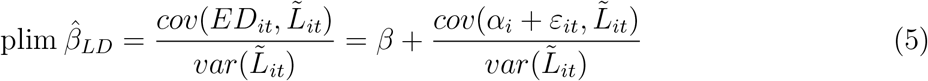

where 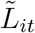 is the residual from the regression of *L*_*it*_ on *ED*_*it−*1_. We use again the definition of 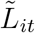 and equation 3 to expand the numerator of equation 5:

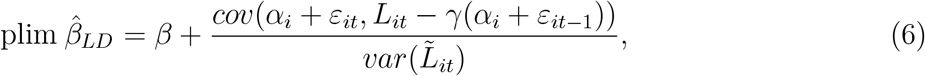

where *γ* is the coefficient of the regression of *L*_*it*_ on *ED*_*it−*1_. Finally, using the expression of *γ* = *cov*(*L*_*it*_, *ED*_*it−*1_)*/var*(*ED*_*it−*1_) and assuming no serial correlation in the error term *ε*, 6 becomes:

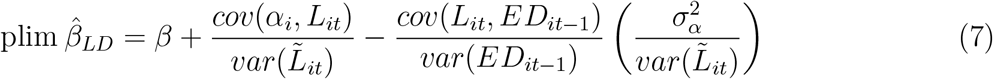

Finally, it is possible to re-arrange 7:

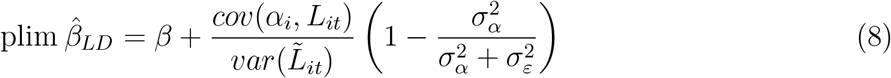

From 8, the bias of 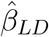 crucially depends on the correlation between *L* and fixed municipalities characteristics *α*_*i*_. If this correlation -as it seems the case in our setting (see Figure 1 in the paper) -is positive, we have that 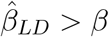 from model 2. Hence, if 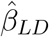 and *β* from model 4 and 2 and are negative, in absolute value 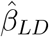 will be lower than *β*. The intuition is the following. Since municipalities with a higher excess mortality are those that are “treated” with *L* = 1 and since fixed differences are positively correlated to *L*, controlling for lagged value only exaggerates the effect of *L*_*it*_ on excess deaths.

Assume, instead, that the correct specification includes the lagged value of excess mortality:

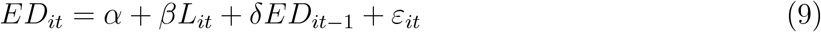

but a regression with fixed effect only is estimated. In this case, by first-differencing 2, we obtain:

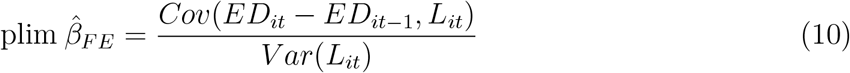

From equation 10 we can substitute *E*_*it*_ *− ED*_*it−*1_ = *α* + *βL*_*it*_ + (*δ −* 1)*ED*_*it−*1_ + *ε*_*it*_ to rewrite 10 as:

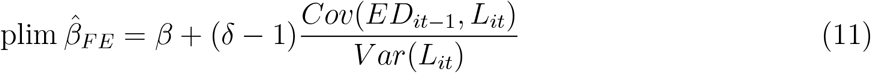

Since in our setting lagged excess deaths *ED*_*it−*1_ and *L*_*it*_ are positively correlated, as far (*δ −* 1) is negative, we have that 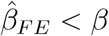 in 10. In our exercise, *δ* is greater than zero and strictly less than 1 as there is some persistence in the stochastic process of *ED*_*it*_.

## Footnotes

1 The Italian Government had also ordered the closing of schools and universities on March, 4.

2 See also Buonanno *et al*. (2020) for the use of excess-deaths as a proxy of Covid-19 deaths for the case of Italy.

3 There are 110 provinces in Italy, with a mean and median population of around 540,000 and 370,000 residents, respectively. The provincial mean and median area is of around 2,700 and 2,400 squared kilometers, respectively. Each province belongs to one of the 20 Italian regions. The public health care system in Italy is managed at the regional level, hence all municipalities within a province are subject to the same rules and have access to the same set of public health facilities.

4 Moreover, official data on daily Covid-19 cases, or deaths classified as being related to Covid-19, are currently only available at the province level in Italy.

5 Note that by “active population” we refer to the number of employed workers (15 years old and above) actively working in a given period (i.e., not employed in sectors subject to the lockdown). This should not be confused with the concept of “activity rate”, which refers to the ratio of the total labor force (i.e., employed and unemployed) to the population of working age. Results are robust to use the labor force population (15 years old and above) as denominator, see Table B.1 in the Online Appendix.

6 Accordingly, we measure the reduction in the share of active population in percentage points rather than as a percentage change. This choice is motivated by the fact that for the diffusion of a virus the level of susceptible individuals matters, as exemplified by the SIR class of models (Kermack and McKendrick, 1927).

7 As shown later, the results are robust to a simple linear specification in which we regress the *ED* on the time-varying share of active population at the municipality-day level. The results are also robust to considering alternative cutoffs (i.e., 25th or 75th percentile in the reduction of the share of active population), see Table B.2 in the Online Appendix.

8 Following Angrist and Pischke (2008) and Guryan (2001), in the Online Appendix D we provide a formal argument of why our approach of estimating 1 may provide bounds to the causal effect of *L* on excess mortality.

9 Table B.3 in Online Appendix B shows that the results are marginally significant for individuals above 40 and, overall, mainly driven by the cohorts above 50.

10 To “depurate” the mobility series from daily patterns, such as drops in mobility on weekends, we use residuals with respect to day-of-the-week fixed effects.

11 The share of active in the first lockdown is instead equal to the overall share of active population, as ISTAT does not provide data on the number of active workers in the first lockdown.

12 INPS-INAPP (2020) shows that the economic sector involved in the second lockdown were the ones where workers are more likely to be at risk of contagion due to proximity and (limited) work-from-home (WFH).

13 Accordingly, we also do not use the variation in the share of active population as an instrument for the differential change in mobility because there are several potential violations of the exclusion restriction. The economic lockdown may affect mortality by Covid-19 through the changes in behavior (for example the use of masks), human interactions (for example spending more time at home), and expectations that are unrelated to mobility. Hence, we see mobility as an important but not unique channel through which the economic lockdown reduced mortality by Covid-19.

14 A detailed chronology of the NPIs, including the regional measures approved before the first nation-wide lockdowns of March 4 and 11, and detailed descriptions of all the measures and legal references, is available on the wikipedia page for the Covid-19 pandemic in Italy.

15 Our deaths data covers the period up to April 30, hence right before the relaxation of the lockdown measures on May 4.

16 This register does not contain information on public sector employees. For further information see: http://dati.istat.it/OECDStat_Metadata/ShowMetadata.ashx?DataSet=DICA_ASIAULP.

17 Notice that a later Decree on March 25 provided a revised list of ATECO sectors. We use this definitive list as a benchmark for the ATECO sectors that were allowed to operate in the second lockdown.

18 Notice that the list of ATECO sectors contained in the Decrees specified active sectors also at the 4 or 5 digits level. As we have information on the number of workers only at the 3 digits level, we consider as active any ATECO 3 digit level embedding the 4 or 5 digits ones. This implies that our measures tend to overestimate the number of active population. Table 3 in the paper provides a robustness check using data on 5 digits level for the second lockdown provided by Istat.

19 As in Figure 2, to “depurate” the movement data from daily patterns, such as drops in mobility on week-ends, we use the residuals with respect to day-of-the-week fixed effects.

20 The data record the most common location of users within a given a time-window, where “location” refers to a *tile*. Tiles correspond to granular locations typically smaller than a municipality. We collapse the data, at the daily frequency and municipality level, distinguishing users moving in or out of a municipality during the day. We, then, compute a “in and out” measure by summing these two variables. Finally, we normalize this measure by computing the number of users moving in and out of a municipality per 1,000 residents in the municipality.

21 For comparability with the Facebook data, we construct province level movements from the municipal EnelX data by taking the average of all municipalities in the province.

22 We construct all mobility indicators as residuals from regressions of the raw data with respect to day-of-the-week fixed effects. The large drop in mobility observed on April 13 corresponds to the 2020 Monday Easter holiday.

